# Pangenome graphs improve the analysis of rare genetic diseases

**DOI:** 10.1101/2023.05.31.23290808

**Authors:** Cristian Groza, Carl Schwendinger-Schreck, Warren A. Cheung, Emily G. Farrow, Isabelle Thiffault, Juniper Lake, William B. Rizzo, Gilad Evrony, Tom Curran, Guillaume Bourque, Tomi Pastinen

**Affiliations:** Quantitative Life Sciences, McGill University, Montréal, QC, Canada; Genomic Medicine Center, Children’s Mercy Hospital and Research Institute, KC, MO, USA; Pacific Biosciences, Menlo Park, CA, USA; Child Health Research Institute, Department of Pediatrics, Nebraska Medical Center, Omaha, NE; Center for Human Genetics and Genomics, Department of Pediatrics, Neuroscience & Physiology, New York University Grossman School of Medicine, New York, USA; Children’s Mercy Research Institute, Kansas City, MO; Canadian Center for Computational Genomics, McGill University, Montréal, QC, Canada; Department of Human Genetics, McGill University, Montréal, QC, Canada; Institute for the Advanced Study of Human Biology (WPI-ASHBi), Kyoto University, Kyoto, Japan; Victor Phillip Dahdaleh Institute of Genomic Medicine at McGill University, Montréal, QC, Canada

## Abstract

Rare DNA alterations that cause heritable diseases are only partially resolvable by clinical next-generation sequencing due to the difficulty of detecting structural variation (SV) in all genomic contexts. Long-read, high fidelity genome sequencing (HiFi-GS) detects SVs against reference genomes with increased sensitivity and also enables the assembly of personal and graph genomes. We leveraged standard reference genomes, publicly available human haploid assemblies (n=94), together with a large collection of HiFi-GS data from a rare disease program (Genomic Answers for Kids, GA4K, n=574 assemblies). These data allowed us to build a deep population graph genome distinguishing very rare SVs from recurrent polymorphisms. Using graphs to discover SVs, we obtained a higher level of reproducibility than that obtained by the standard reference approach. We observed over 200,000 SV alleles unique to the rare disease GA4K cohort, including nearly 1,000 rare variants that impact coding sequence. With improved specificity for rare SVs, we isolate 30 candidate SVs in phenotypically prioritized genes, including known disease SVs. We isolate novel diagnostic SV in *KMT2E* in a patient demonstrating use of personal assemblies coupled with pangenome graphs as a new handle for rare disease genomics.

## Introduction

Structural variants (SVs) contribute to Mendelian and complex disease, yet they are the most challenging to detect, assemble and fully resolve. Indeed, many SVs are in repetitive sequences that are difficult to approach with short-read sequencing libraries ***(1, 2)***. In contrast, long-reads can detect and characterize many more complex and repetitive structural variants ***(3)*** and can be used to efficiently build reference-free *de novo* assemblies from genomes as large as a human genome ***(4)***. Long-reads together with information from parental sequencing also enable the phased assembly of haplotype resolved maternal and paternal genomes based on unique k-mers ***(5)***. Previous efforts employing long-reads have discovered up to 28,000 SVs per human genome ***(6)***. However, we still need to adopt computational methods to fully leverage this richer data in the context of rare disease. The traditional approach of comparing a proband genome against a single reference genome may fail, even with high quality genome assemblies, since some regions may be absent or present very different alleles. Moreover, it remains difficult to compare alleles between genomes, since the genomes are only related to the reference genome and not to each other. While tools exist to call and cluster SVs ***(7-9)***, they rely on heuristics such as the maximum distance between events, which may erroneously split or merge SVs because they do not consider the entire genome assembly. Therefore, accurate estimation of SV allele frequency still requires better tools.

To alleviate these shortcomings and fully leverage high quality genome assemblies, a pangenomic approach, where genomes are related to each other in a graph, is necessary. Some approaches have been developed to achieve this through progressive alignment of genome assemblies ***(10, 11)***, while others do so through pairwise comparisons ***(12)***. The resulting pangenome graphs, built from high quality genome assemblies, have already been used to create a pangenome reference of human population diversity by the Human Pangenome Reference Consortium (HPRC) ***(13)***. This new type of reference showed increased sensitivity in detecting SVs over methods that use a linear reference. Pangenome graphs also have the potential to provide more robust allele frequencies, especially in the case of multiallelic SVs. Here, we explore the benefits of using such a strategy to discover rare alleles in a disease cohort. Specifically, we show how pangenome methods can be used along other tools to improve sensitivity and specificity in detecting rare SVs.

## Results

### A pangenomic approach to identify and integrate structural variation across hundreds of genomes

We pursued discovery of rare SVs that were potentially pathogenic among a cohort of 287 parent-offspring trios included in the Genomic Answers for Kids (GA4K) program targeting pediatric genetic disease ***(14)***. By design, this cohort was enriched for difficult to solve cases and more than 90% of the probands were undiagnosed even after standard clinical sequencing and exploration of putatively causative single nucleotide variants (SNVs). We included short-read genome sequencing (srGS) parental data and further sequenced all the probands using PacBio HiFi reads (Methods). A subset of this HiFi-GS data, but none of the assemblies, was included in an earlier study ***(14)***. Here, we expanded the cohort and systematically developed assemblies of 574 haploid proband genomes using hifiasm ***(15)***. To facilitate the identification of rare variants, we also augmented our data with the 94 haploid genomes released by the HPRC ***(13)***. We then created a pangenome graph with minigraph ***(10)*** to identify structural variants in the combined set of 668 haploid genomes together with two standard reference genomes (GRCh38 and chm13v2). We chose minigraph since it scales linearly with the number of genomes at the expense of requiring a backbone reference to build the graph. During graph construction, when a haploid genome was added, polymorphisms that were larger than 50 base pairs (bp) but missing from the graph created new nodes and paths (see **Fig 1A**). With our data, we found that the number of new non-reference sequence nodes added from each additional haploid genome plateaued at around 500 (**Fig 1B**). This suggests that there remains many more alleles to be discovered in human genomes, continuing the trend previously observed in the HPRC dataset ***(13)***.

**Figure 1:**
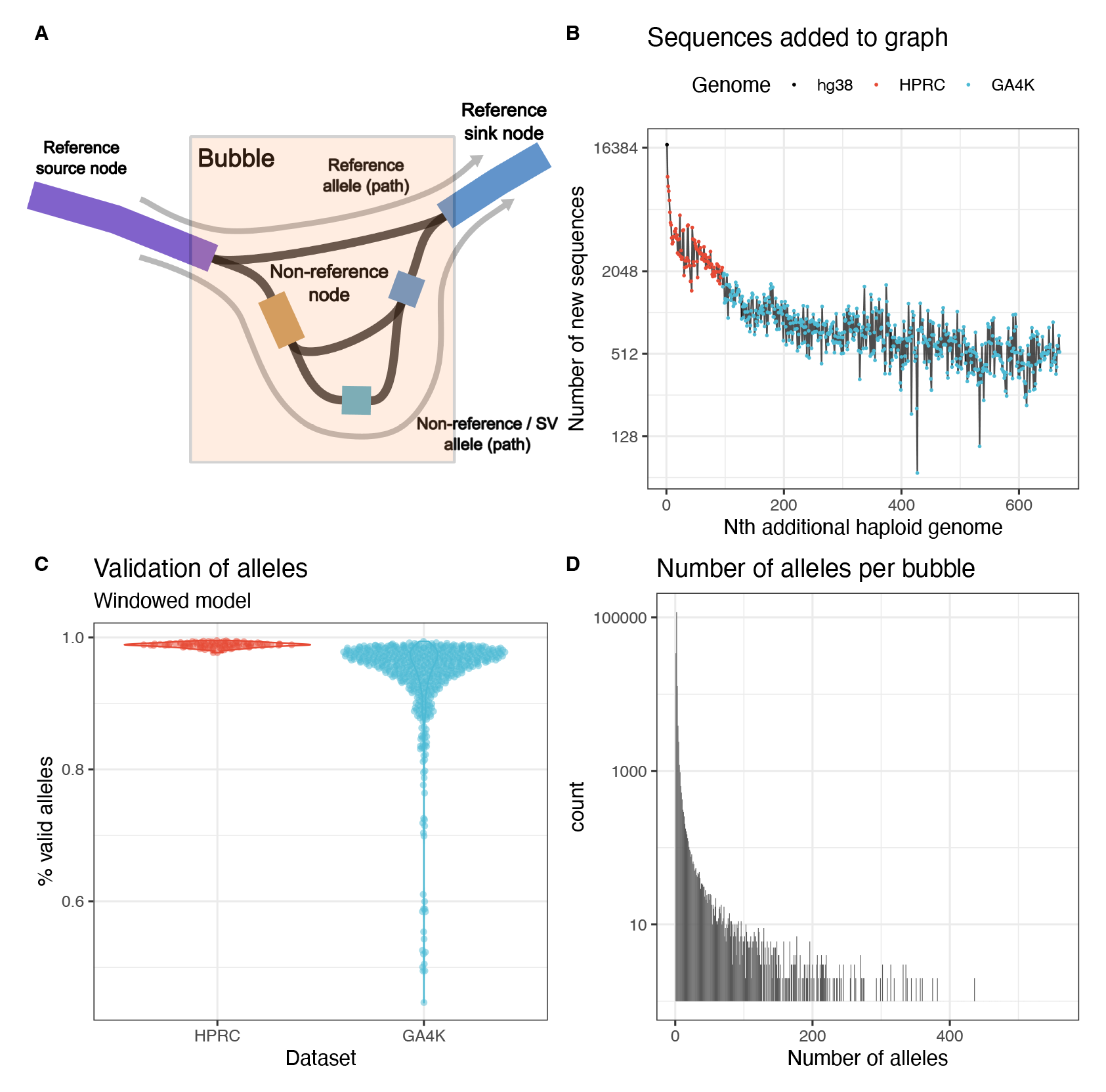
Construction of the pangenome. A) Representation of a polymorphic locus in a genome graph. A bubble begins at a source node when at least two genomes are different and ends at the sink node, where all genomes are the same. Paths from the source node to the sink node are alleles. Non-reference nodes are new sequences found in one of the 668 assemblies. B) Growth of the pangenome as new genomes are added. C) Proportion of genotypes supported by sequences that pass validation with Flagger. D) Distribution of the number of alleles observed in each bubble among the 668 genomes.

Using the resulting graph we genotyped the assemblies and observed 180,755 bubbles and 631,400 distinct alleles (see **Fig 1A**). To ensure all genotypes were derived from reliably assembled sequences, we validated the assemblies with Flagger ***(13)*** and excluded the genotypes supported by collapsed, duplicated or low coverage regions (Methods). In the best assemblies, 98% of alleles were supported by valid regions, which is comparable to the HPRC assemblies (**Fig 1C**). In a subset of 43 lower coverage assemblies, the number of alleles supported by valid regions was as low as 45% (**Fig S1**). However, Flagger may not be suitable to validate lower coverage assemblies where the sample size in each region is too low because it needs to fit a mixture model on genome coverage data ***(13)***. As expected, singleton alleles were the most likely to be called from misassembled sequences, while very common alleles were the least likely (**Fig S2**). After excluding genotypes from invalid regions, we count a total of 178,188 reliable bubbles, involving 501,967 non-reference nodes and constituting 584,146 alleles. Some of these alleles were found in biallelic loci (150,942 alleles) but most were found in complex polymorphic loci (433,204 alleles) where we count up to 560 alleles in the same locus (**Fig 1D**). Ancestry mapping based on SNVs (**Fig S3**) and pangenome alleles (**Fig S4, S5**) showed that the majority of GA4K probands are of European-ancestry (EUR) and in line with self-reported ethnicity characteristics of the GA4K cohort ***(14)*** (Methods).

### Repeats and duplications are major contributors to structural variation

The non-reference nodes in the pangenome graph contributed approximately 610 million base pairs (Mbp) of non-reference sequence, of which 184 Mbp was derived from HPRC genomes and 426 Mbp from GA4K genomes. We wanted to know what contributes to this increase in the size of the pangenome. RepeatMasker found that 74.2% of the content in non-reference nodes and that the leading contributors were simple repeats, satellites, L1s and Alus (**Fig 2A**). In terms of alleles, the pangenome contained 57,129 insertions, 50,011 deletions and an additional 418,302 variants with paths that pass through complex bubbles. These complex bubbles represent multiple deletions, insertions or substitutions of DNA segments in loci such as STRs or VNTRs where structural variation from multiple genomes overlaps. Moreover, we discovered 1,056 full-length LINE polymorphisms (Methods) and 15,598 full-length SINE polymorphisms (**Fig 2B** and **S6**), of which 340 LINEs and 3,664 SINEs are unique to GA4K (**Fig S7**).

**Figure 2:**
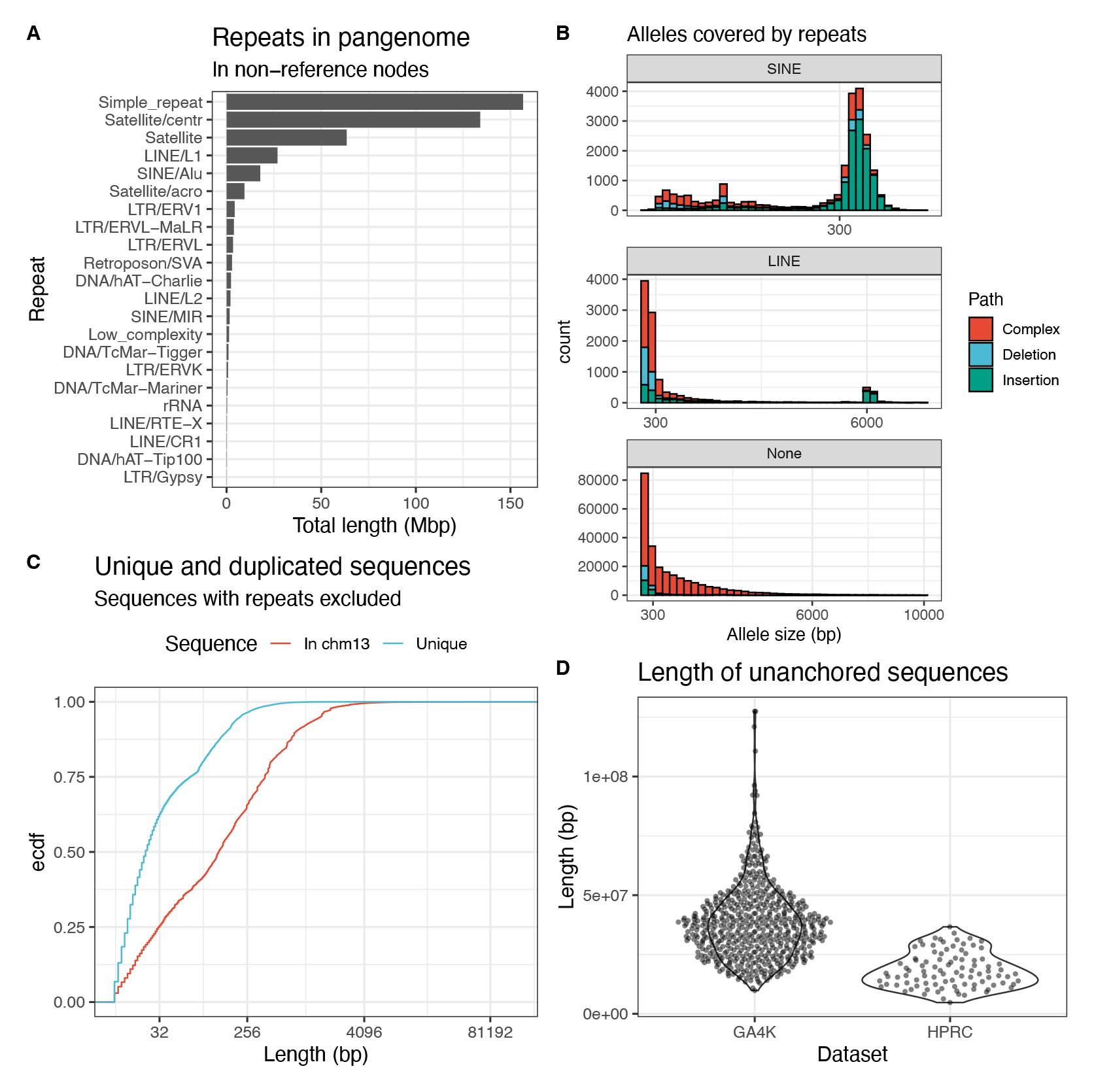
Contents of the pangenome. A) Length of pangenome (non-reference graph nodes) that is masked with RepeatMasker. B) Length distri-bution of alleles that are covered by SINE and LINE sequences, and alleles that are not covered by repeats. C) Length distribution of sequences that map to chm13v2.0 but are not repeats and of sequences that do not map to chm13v2.0 and are not repeats. D) Total length of sequence in each sample that is not anchored in the pangenome.

Next, we aligned the non-reference nodes with no repeats back to the CHM13v2 reference and found that 24.5% of all non-reference sequences (8.9% in HPRC and 15.6% in GA4K-only) mapped to some region in the genome (**Fig 2C**), suggesting duplication or other rearrangement events. This left 1.3% (7.5 Mbp) of the pangenome as putatively novel sequences that did not align to repeat databases or the reference genome. Novel sequence from the GA4K proband genomes accounted for 0.8% (4.7 Mbp) of the pangenome. Many of these novel sequences assigned to the pangenome were short, with only 21% being longer than 100 bp (**Fig 2C**).

Finally, separate from the non-reference nodes that were included in the graph, there are also unanchored contigs in both HPRC and GA4K assemblies (**Fig 2D**). We independently confirmed some of the unanchored contigs for one offspring via coverage from mapped srWGS sequencing reads from that sample and its parents, and validate that contigs inherited from one parent (where there was high coverage from the offspring and only one parent) were unambiguously almost always inherited only from the appropriate parent, (**Table S1**), indicating that these unplaced portions of the pangenome graph can be additional, potentially *de novo*, family-inherited DNA sequence. We also confirm RNA transcription from these srWGS validated haplotype-resolved unanchored contigs by the mapping of phased reads from IsoSeq sequencing of the offspring (**Fig S8**), showing that these sequences could potentially contain active genes not currently captured in the reference genomes. In these unanchored contigs that are not in the graph, an average of 69 kilo base pairs (kbp) of sequence per genome did not contain repeats and did not align to Chm13v2 (**Fig S9**).

### Calling SVs with a pangenome graph improves error rates

We expect pangenome graphs to recover the SV alleles called by other long-read methods. We calculated the recall and precision of minigraph SVs (non-reference alleles in the genome graph) over the 287 probands against the SVs obtained with PBSV (Methods), which uses unassembled PacBio HiFi reads aligned to the GRCh38 reference genome. This yields a two dimensional distribution describing the recall and precision for each minigraph SV in each sample, which we visualize as a heatmap (**Fig 3A**). We note that most SVs achieve very high precision and recall, while a small number show lower precision or lower recall. Over all genotypes, minigraph achieves a recall 0.78 of and a precision of 0.80 against PBSV. Since no truth set is available, we could not directly evaluate the true positive rates of minigraph and PBSV. However, there is a twin pair in the GA4K cohort that we can use to explore the rate of SVs that replicate as a proxy for the true positive rate. We found that PBSV calls a total of 23,516 SVs, of which 19,547 (83.12%) are replicated in both twins (**Fig 3B**). Meanwhile, minigraph calls 29,964 SVs, of which 25,456 are in both twins (84.96%). This boost in the number of detected SVs is corroborated by previous findings showing an increase in sensitivity over reference-based methods ***(13)***. These results also indicate that false positive and negative rates of PBSV and minigraph heavily impact allele sharing since 15.04% (minigraph) and 16.88% (PBSV) of alleles were detected in only one of the twins. Thus, we consider increased allele sharing between siblings to be evidence of lower false positive and negative rates. We then explored allele sharing within the other 58 GA4K families in which at least two siblings were sequenced. In high quality pairs where both siblings were sequenced at a depth above 20X minigraph shows an average of 7.1% more allele sharing than PBSV (**Fig 3C**). As expected, lower coverage samples show less allele sharing due to their higher error rates (**Fig S10**). If we include pairs sequenced at a lower depth, siblings share on average 3.3% more alleles with minigraph than with PBSV (**Fig S11**). The increase in allele sharing between siblings suggests that the SV calls obtained with minigraph have a lower false positive and false negative rate.

**Figure 3:**
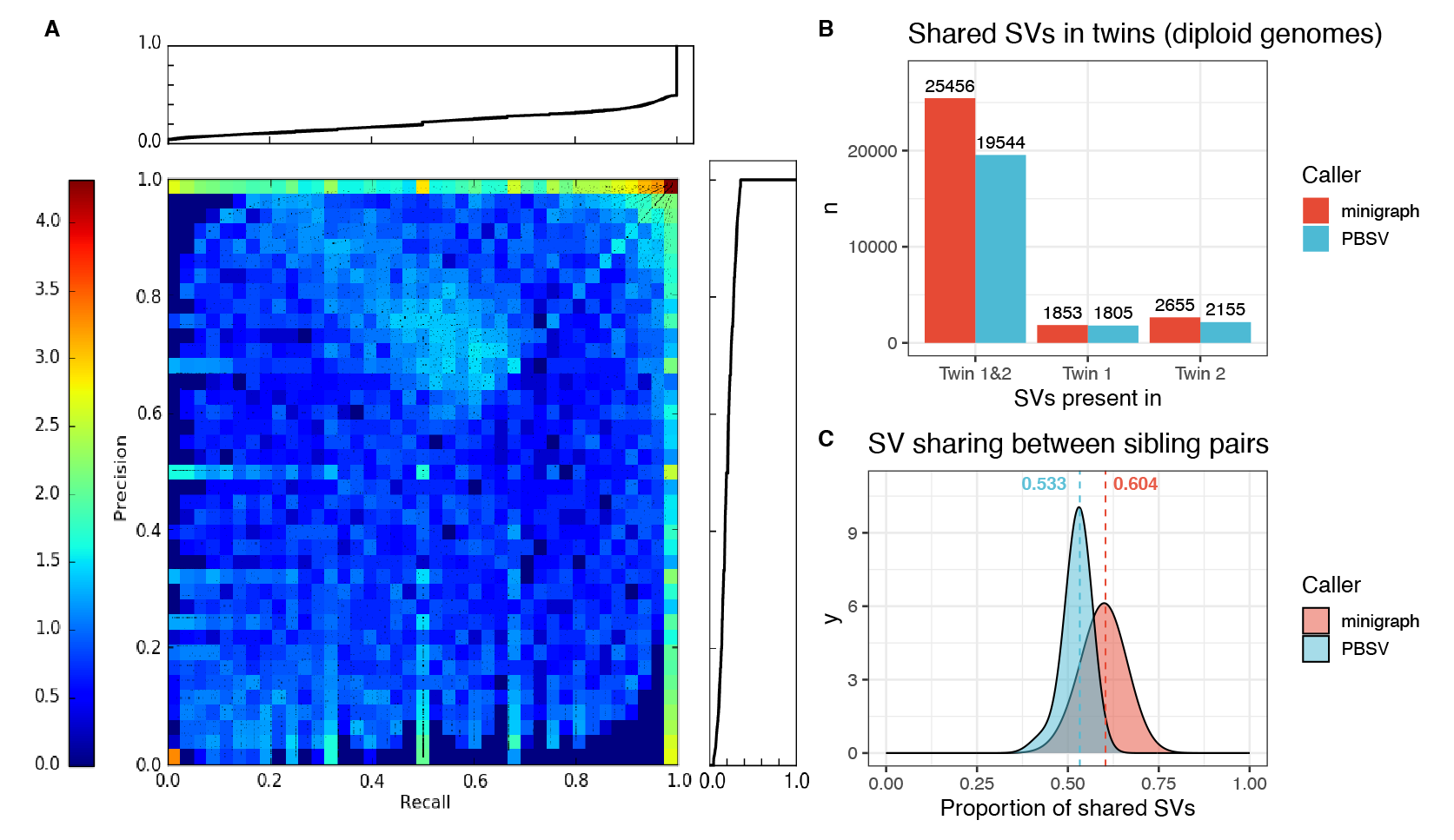
Validation of SV calls. A) Recall and precisions of minigraph SV calls against PBSV. B) Allele sharing between the GA4K twin pair, calculated with PBSV versus minigraph. C) Allele sharing between siblings in GA4K (twins and low coverage pairs excluded), calculated with PBSV ver-sus minigraph.

### Pangenome graphs reveal rare SV alleles in haplotype resolved assemblies

On average, we genotyped 18,326 non-reference SVs per sample (**Fig S12**), a figure that is in line with previous findings ***(6)*** but that is also influenced by assembly quality (**Fig S13**) and genome diversity. We wanted to characterize the population frequency of alleles in this dataset to identify rare variants in our set of individuals. To achieve this, we split alleles into three groups: those that are common to both cohorts, those that are unique to HPRC and those that are unique to GA4K. As expected, the majority of alleles were observed in both cohorts, and their frequency distribution features the full range of rare, common and nearly fixed alleles (**Fig 4A**). More precisely, we observed 185,926 singleton alleles, 389,983 alleles with a frequency below 10% and 66,034 alleles with a frequency above 90%. In total, 314,981 alleles were observed in both datasets, 64,614 were unique to HPRC and 204,551 were unique to GA4K. The most common allele unique to GA4K occurs in 88 out of 574 haplotypes (**Fig 4A** bottom). Similarly, the most common allele unique to HPRC occurs in only 23 out of 90 haplotypes. As expected, the allele frequency distribution of alleles that are unique to GA4K and HPRC is heavily skewed toward rare variants that occur with a frequency below 10% (**Fig 4B**). Next, to begin exploring the properties of rare alleles, we categorized alleles into insertions, deletions and complex events that may not be simple insertions or deletions relative to the reference and checked their size. We found that the average allele was 8.7 kbp long, with the expected peak at 300 bp (corresponding to Alu-related events, and that some alleles could reach up to 100 kbp in length (**Fig 4C**). Notably, rare SVs were found to be longer than common SVs.

**Figure 4:**
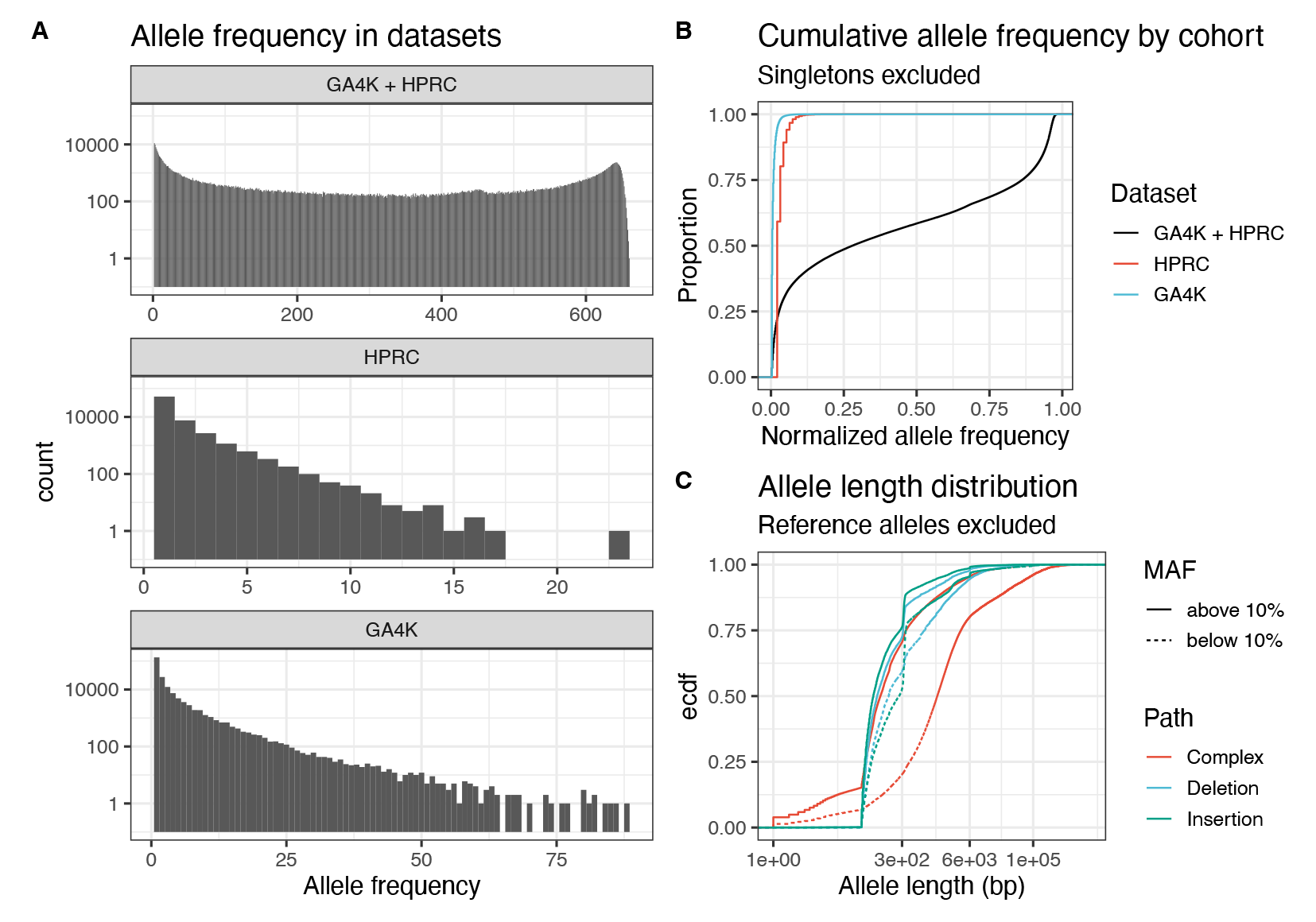
Frequency of SVs. A) Frequency distribution of alleles that are found in HPRC and GA4K, HPRC-only, and GA4K-only. B) Cumulative distribution of allele frequencies (scaled by the size of HPRC and/or GA4K) in these three subsets of alleles. HPRC-only and GA4K-only variants are skewed towards rare frequencies. C) Length distribution of non-reference SV alleles, stratified by frequency and type.

### Rare SV alleles are distributed across the genome and found in genes of interest

Next, we were interested in SV alleles that may have functional relevance. To this end, we focused on the 204,551 alleles that were unique to GA4K and of which 132,391 SVs are singletons. We observed that these alleles occur in hotspots of structural variation near telomeres and centromeres and that they sometimes overlap with genes and exons (**Fig 5A**). Overall, we found 73,982 alleles within genes (**Fig S14**), of which 18,095 were within exons (**Fig 5B**). In particular, 1,383 alleles overlap 306 OMIM ***(16)*** exons that were previously associated with Mendelian diseases and phenotypes (**Fig 5C**). When binning these alleles by frequency, the majority were singletons and rare variants. Singleton SVs accounted for 51,733 SVs in genes, 13,083 in exons and 978 in OMIM exons. As expected, the frequency spectrum in exons and OMIM exons showed the strongest skew towards rare alleles, followed by intra-genic regions and intergenic regions (**Fig 5D**). In particular, 72.4% of SVs in exons and 70.7% of SVs in OMIM exons were singletons. In comparison, singleton SVs were slightly less common in genic regions (69.2%) and much less common in intergenic regions (62.7%).

**Figure 5:**
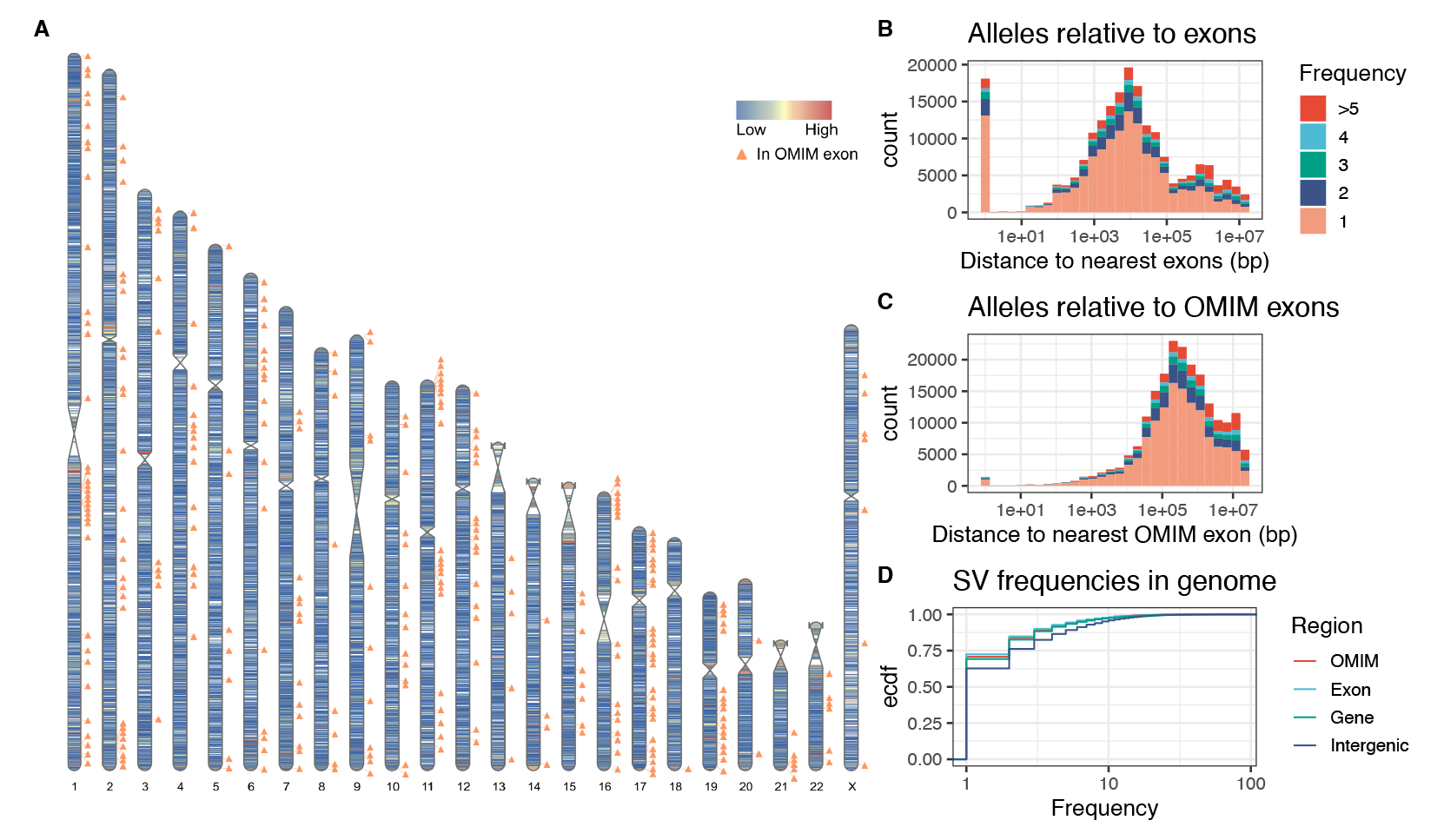
Distribution of rare SVs in the genome. A) Density and hotspots of rare SVs that are found only in the GA4K cohort. OMIM exon positions are annotated. B) C) Distance distribution of GA4K-only SVs relative to exons and OMIM exons stratified by frequency. D) Allele frequency spectra of SVs in intergenic, genic, exonic and OMIM regions of the genome.

### Improved rare variant calling by joint graph and reference-based approaches

We have noted previously that rare HiFi-GS SV variants have higher parental transmission than rare short-read SV calls ***(17)***. However, both pangenome and reference methods still display substantial false positive rates for rare SVs, since every false positive will occur only a few times and be mistaken for a rare SV. We hypothesized that a consensus of reference-based and assembly-based methods would improve the precision of rare SVs over reference-based methods alone. To test if the proportion of false positives relative to true positives was reduced by such an approach, we created a consensus set of SVs that were called with both minigraph and PBSV. We then separated the PBSV calls into two sets: those that overlap minigraph SVs (concordant set) and those that do not overlap minigraph (discordant set). We did this for all calls, and again only for rare SVs with a frequency below 5%. Then we checked how many SVs in the test set were reproduced by an independent set of Illumina srGS Manta ***(18)*** SV calls (Methods). For all SVs, we found that the srGS SVs reproduce 56% of the concordant set and 33% of the discordant set for all calls. In the case of rare SVs, Illumina Manta SVs reproduce 61% of concordant SVs and only 8% of discordant SVs (**Table S2**), indicating that the false positive rate is indeed lower within the minigraph and PBSV concordant calls. Thus, combining assembly and reference-based methods improves the precision of rare SV calls significantly.

### Discovery of phenotypically impactful structural variants

Finally, using this strategy, we curated the GA4K-specific minigraph alleles (not observed in HPRC) that are replicated by PBSV and are potentially disrupting exonic sequences (n = 924). To focus on variants with potential phenotypic impact, we used the patient structured phenotype terms (HPO) to score candidate loci for each patient and limited to the top quartile of scores (phrank > 5) ***(19)***. Among the rare SVs impacting the resulting 40 filtered exons, 10 were seen in highly polymorphic (non-constrained) exons or mapped to non-OMIM genes. In the remaining 30 exons, we observed 23 potentially pathogenic disruptions (2 previously reported pathogenic in ClinVar) in genes where loss-of-function (LOF) alleles are reported causes, but for autosomal recessive diseases (**Table S3**). We checked for additional possible causal SNVs among the individuals with these 23 potentially deleterious alleles using DeepVariant ***(20)*** and did not find any. In three other cases, the nature of the variant and its inheritance from the unaffected parent suggested low disease relevance. However, a subset of four alleles included a previously detected, causal structural variant in *AARS2* (**Fig S15**) where the second variant is a SNV in *trans* (data not shown). Also, a disease candidate inversion involving *ACOX1* locus was observed, however typically dominant *ACOX1* mutations are gain-of-function, and therefore follow-up RNA expression studies are required (**Fig S16**). A paternally inherited rare deletion in *NLRP12* was observed with partial phenotypic fit, where variants have reported to have variable penetrance. Finally, a novel diagnostic finding was uncovered in the maternal haplotype of one patient: a 14.5kb deletion in *KMT2E* ranking in the top 5^th^ percentile in phenotype fit score (phrank) ***(19)*** among all disease genes in this proband and was the highest scoring exonic rare SV affecting exons 9-13 in *KMT2E* (**Fig 6A-C**). This deletion is predicted to result in a premature stop and loss of function (NM_182931.3(*KMT2E*):c.729+113_1359-612del (p.Ala244*). The patient had a neurodevelopmental phenotype of hypotonia, macrocephaly and developmental delay, overlapping the clinical picture described for *KMT2E* loss-of-function (AD) variants. The maternal transmission was verified from sequence reads (**Fig 6D**), the variant was validated by short-read genome data, and was clinically confirmed by long range PCR. Importantly, the mother had a history of cognitive delay and learning disabilities.

**Figure 6:**
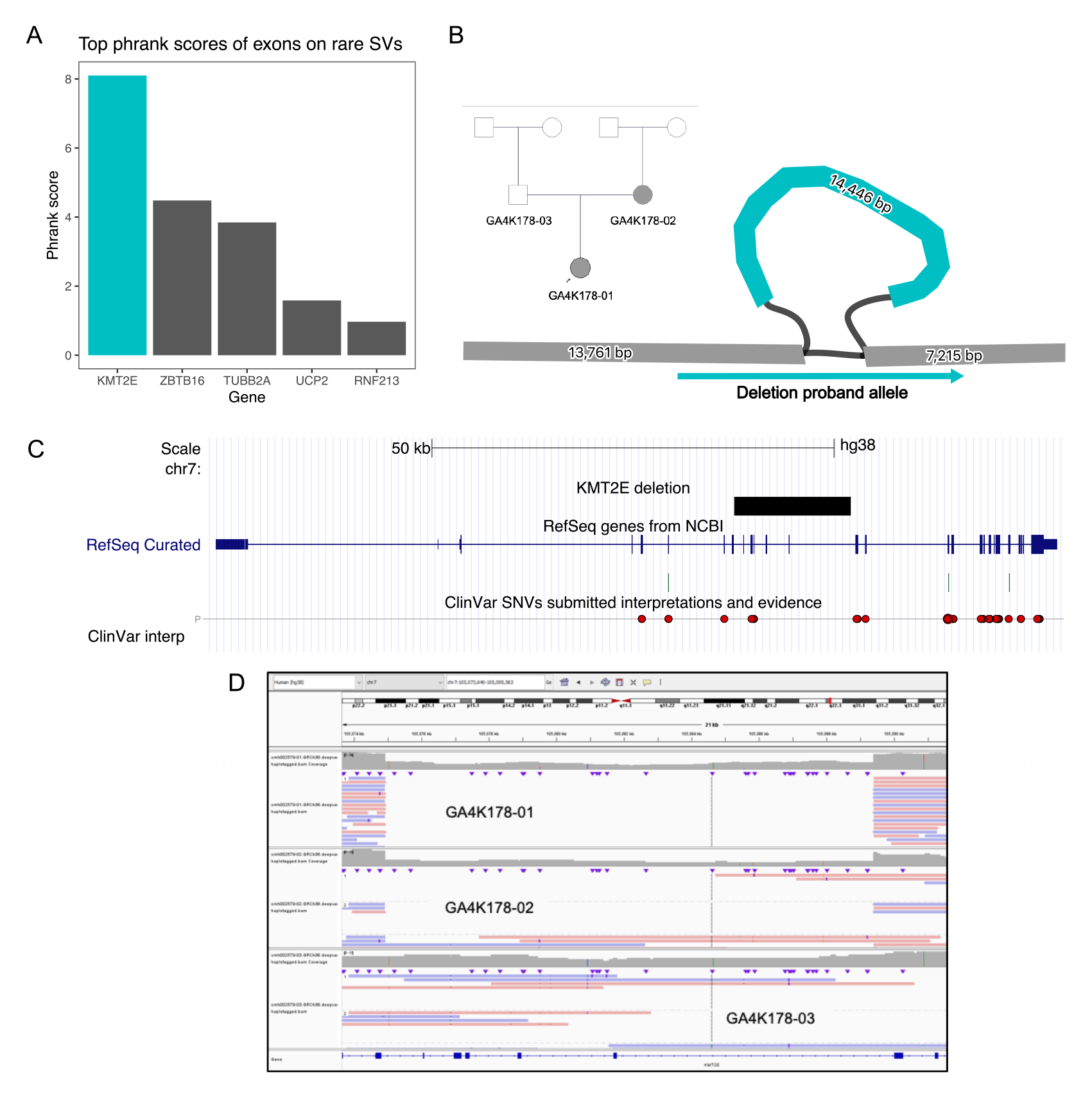
*KMT2E* diagnostic deletion. B) Phrank rankings of rare SVs within exons. The *KMT2E* rare deletion ranks first among all rare SVs that overlap exons. B) Pedigree of the 14,446 bp deletion, and its representation in the genome graph. C) UCSC Genome Browser view of the affected region. D) Raw alignments of long-reads in the proband genome, the maternal and the paternal genomes, confirming the deletion.

## Discussion

Pangenome graphs provide a comprehensive framework to study genetic variation and can explore complex loci that are difficult to characterize from pairwise comparisons to a reference genome. The ability of genome graphs to resolve recurrent structural variant biology ***(21)*** and repetitive DNA ***(22)*** was highlighted following the completion of the draft human pangenome reference ***(13)***. In the case of rare genetic diseases, causal variants have a population frequency that is significantly lower than 1%. Expanding the collections of personal haploid assemblies in each population and generating deeper pangenome graphs will improve the filtering of common and rare SVs and help the identification of ultra-rare genetic variation in proband genomes.

Here we relied on a progressive pangenome construction technique where each proband genome was added one at a time revealing the SVs that are common and those that are unique. Such a progressive method is efficient but a limitation is that it depends on the order of genomes that are incorporated and might miss events such as translocations ***(23)***. An alternative could be to use a reference-free method such as the PanGenome Graph Builder ***(12)*** but further developments would be needed to implement adding genomes to an existing pangenome.

Most of the additional pangenome content is composed by repeats, genomic duplications and other rearrangements, which are difficult sequences that sometimes exceed even the length of long reads. This is especially the case for rarer SVs, which tend to be longer. These limitations increase the genotyping error rate which complicate the filtering and ranking of pathogenic SVs. Ensemble approaches that combine orthogonal approaches have been shown to improve variant calling ***(24-26)***. Our analyses combined graph and reference-based approaches to improve the accuracy of SV identification. We also applied phenotypically guided prioritization for manual curation involving only coding structural variation in known disease genes. Therefore, other novel disease genes and potentially non-coding variation can remain in our dataset and may be important for unsolved cases.

Finally, the rare alleles represented in the pangenome graph do not include sequences that could not be anchored to the pangenome. The estimated 69kb of unique sequences in contigs outside the pangenome were corroborated by family inheritance patterns and showed evidence of transcription. Placement of these contigs might require greater read lengths and sequencing depths, but further experiments would be needed to understand their potential function.

## Methods

### Trio assemblies with parental HiFi reads

To produce trio-binned assemblies, k-mer hash tables including unique k-mers were created for each parent from HiFi reads using yak count v0.1 (Cheng et al., 2021). Proband HiFi reads were then assembled into two haplotypes with hifiasm v0.15 ***(5)*** using the trio-binning method. The hifiasm k-mer frequency settings were adjusted to have a lower and upper bound of 1 to improve binning at lower parental coverage.

### Trio assemblies with parental short-reads

K-mer hash tables excluding unique k-mers were created for each parent from Illumina reads using yak count v0.1 ***(5)***. Proband HiFi reads were then assembled into two haplotypes with hifiasm v0.15 using the trio binning method.

### Validation of diploid assemblies

We validated the diploid genome assembly using the Secphase-Flagger pipeline ***(13)***. The HiFi reads were realigned to the combined diploid genome of each sample with minimap2 ***(27)***. The alignments were phased, corrected and filtered using Secphase. For correcting alignments, we called biallelic SNVs with DeepVariant on the phased alignments. Then we calculated the coverage across the diploid assembly with samtools depth. Finally, we clustered the regions of the diploid assembly into haploid, error, collapsed and duplicated categories using Flagger. Specifically, we fitted the clustering model over 5 Mbp windows of the diploid assembly to account for local biases in sequencing depth.

### Ancestry mapping

We use somalier ***(28)*** to predict ancestry from the genotypes using principal components analysis, based on 17 766 informative sites and 2504 reference samples from the 1000 Genomes Project.

### Creating genome graphs

We used minigraph ***(10)*** with base-level alignments to build the genome graph. We started with the CHM13v2 ***(4)*** reference as a backbone, and progressively augmented the graph with the hg38 reference, the HRPC genomes, and finally, the 574 GA4K haploid genomes. The order of the genomes to be added to the graph was determined lexicographically by sample name.

### Surveying additional sequences in the graph

We selected non-reference nodes that are above 100 bp in length from the genome graph. We ran RepeatMasker with the Dfam_2.0 ***(29)*** database on the nodes to identify repeats. These nodes were also aligned to CHM13v2 with minimap -x sr to count how much sequence is not observed in the reference genome. Then we removed node intervals covered by RepeatMasker or CHM13v2 alignments with GenomicRanges::subtract to find unique sequences. Unique sequences shorter than 10 bp were ignored. We also spelled the path sequences and RepeatMasked full length alleles.

### Calling genotypes

We called genotypes by realigning the assemblies back to the graph with minigraph -xasm --call. The resulting genotypes were corrected by keeping calls derived from regions labeled as haploid. Calls derived from error, collapsed or duplicated regions were marked as invalid. We repeated the same with HPRC genomes. We merged the genotypes over the entire cohort by enumerating each observed traversal of a bubble as an allele in each sample. Then we created a matrix where alleles are rows and columns are samples and where the presence of an allele is marked with 1 and its absence with 0.

### Annotating alleles

We categorize alleles according to the structure of the bubble they are found in. Some alleles are simple paths that are clearly insertions or deletions, while some are complex paths that may be a combination of insertions, deletions and alternate haplotypes relative to the reference. We ran RepeatMasker over the sequence of each allele. Full length SINE and LINE were defined as alleles that are more than 80% covered by the repeat annotation, and are 250 to 400 bp and 5000 to 10000 bp in length respectively. Each allele was overlapped with genes and exons using the lifted EBI GENCODEv38 r2 ***(30)*** annotation that was published with the CHM13v2 genome ***(4)***. Similarly, alleles were overlapped with OMIM exons using annotation that was lifted over to CHM13v2.

### Population structure from SV genotypes

To compute the population structure of our datasets and the HPRC samples, we selected alleles that are called from a common set of regions that were assembled and passed quality checking in all samples. We augmented the HPRC genomes with 100 GA4K genomes of known EUR ancestry from previous ancestry mapping with somalier ***(28)*** in order to create a training set. Then we used the R prcomp function followed by UMAP ***(31, 32)*** on the first 45 PCs ***(32)*** on this training set to learn a projection, which we then applied to all HPRC and GA4K genomes.

### Comparison of SV genotypes from minigraph with SV calls by PBSV on GRCh38

In order to evaluate the accuracy of our method, we compared minigraph calls to long-read PBSV PacBio calls. For this comparison, we restricted the minigraph calls to those with only two alleles and classified the less common variant as the ALT allele. We further omitted minigraph calls that are less than 100bp away from regions missing in GRCh38. For this comparison, we considered both: (1) the set of all minigraph calls remaining after filtering and (2) only minigraph calls whose ALT allele corresponds to a star (*) deletion. To compare the resulting call sets to PBSV, we then found all (> 50 bp) PBSV calls within 100 bp of a minigraph call. We then quantified the correspondence between minigraph and PBSV by treating PBSV calls as the ground truth and calculating recall and precision accordingly:

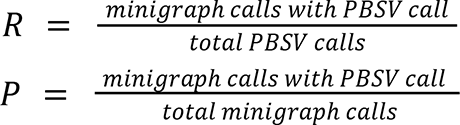

When reporting the average values *R* and *P*, we restricted to regions with at least one minigraph call and one PBSV call in at least one sample.

When analyzing the shared genotyped SVs in pairs of siblings, we used SURVIVOR ***(7)*** to merge the non-reference PBSV calls of the siblings that are at most 10% of the SV length apart. Similarly, we merged the non-reference minigraph alleles delimited by the same source and sink nodes of the SV bubble. Since the PBSV genotypes are not phased, we only considered presence and absence of SV calls. The expected allele sharing is not necessarily 50% due to population structure among parents. To plot the allele sharing density, we adjusted the bandwidth parameter in the density kernel to smooth out the lower modes that are related to differences in sequencing coverage and to emphasize the highest modes that are related to PBSV and minigraph performance.

### Sensitivity of minigraph and PBSV consensus

To combine minigraph and PacBio PBSV calls into a single high-quality dataset and quantify the sensitivity of this set, we separated PacBio calls into those that match minigraph calls (the "concordant" set) and those that do not match minigraph calls (the "discordant" set) and then compared the rates at which the Illumina Manta calls ***(18)*** recall the concordant and discordant set. We used this to measure how the proportion of false positives changes relative to the proportion of true positives in the concordant and discordant sets. For this analysis, we focused on a set of 68 samples that have high-coverage PacBio sequencing data, high-coverage Illumina sequencing data, and minigraph calls. The reported numbers are averaged over these 68 samples. To create the concordant/discordant datasets, we first filtered the minigraph calls to regions consisting of exactly two alleles. We next created two sets of the minigraph/PBSV/Illumina calls - (1) sets containing all calls that pass quality filters and (2) sets containing only rare calls at < 5% MAF. Then, for all and rare datasets separately, we found the PBSV calls that do/don’t overlap a minigraph call to define the concordant/discordant datasets and then determined the fraction of these datasets whose calls overlap an Illumina Manta call.

### Unanchored assembly contigs

For all assemblies, we extracted contigs that do not align to the pangenome end to end. We aligned the sequences with minimap2 to chm13v2.0 to find subsequences that align to the human genome. Then we ran RepeatMasker to identify repeats in these contigs. We reported the amount of unique sequence in each assembly as the number of basepairs that does not align to the pangenome, does not align to chm13v2.0 and is not covered by RepeatMasker annotations.

For two trios where corresponding srWGS data was available, we used the assembled contigs for each of the two phased haplotypes in the child as a personal reference genomes, and used DRAGEN to align the srWGS from each of the parents and from the child itself to the child’s personal reference genomes. Examining the coverage in the assembled contigs that were completely unanchored by the minigraph at 500bp bin resolution using mosdepth, we classified the bins as either being covered (> 8X for the child or father of GA4K86-01, > 3X for other parents), otherwise classified as uncovered. While the majority of bins in the unanchored regions are covered by the all three members of the trio, there is a distinctive subset of over 1Mb of sequence that is covered in the child and the expected parent for the phased haplotype for the personal reference, indicating there is still unique, inherited genomic sequence yet to be explored (Table S1).

For one of these trios, in addition to the srWGS data, we also have Iso-Seq RNA expression for three different cell types (blood, iPSC, neuronally-differentiated iPSC), which we aligned to the paternal and maternal personal genomes of the proband. The majority of the Iso-Seq expression was seen in the bins that are covered by both parents (as that is the largest proportion of the bins), however, among the bins that were uniquely covered by only one of the parents, we see Iso-Seq signal aligned to the paternally inherited genome only showed expression in bins covered by the father, and likewise Iso-Seq signal aligned to the maternally inherited genome only showed expression in bins covered by the mother (Fig S6).

## Data Availability

Data are accessible through DbGaP controlled access.

## Pangenome graphs to improve the analysis of rare genetic diseases: Supplementary data

## Supplementary Tables

**Figure S1:**
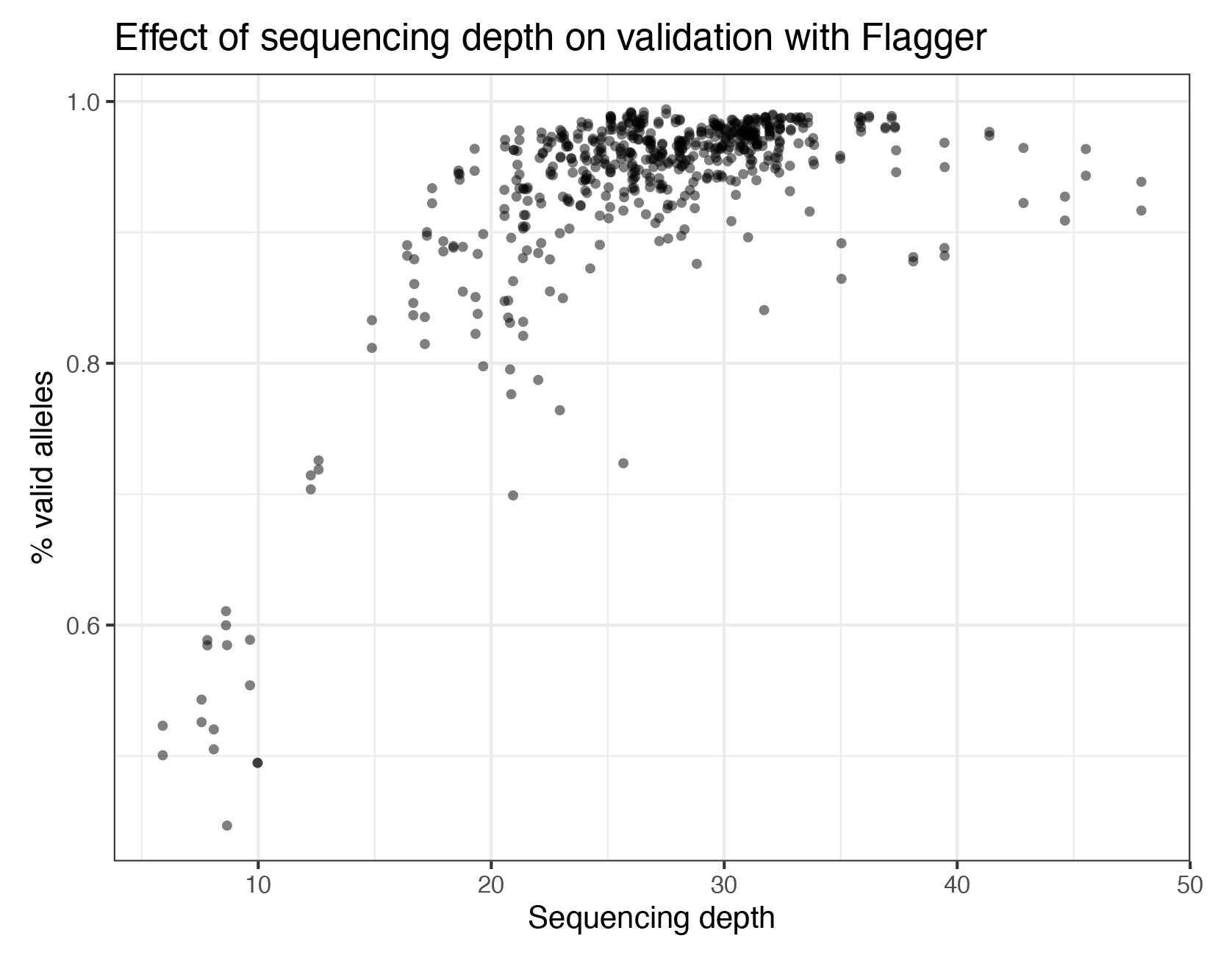
The proportion of alleles that pass validation with Flagger in a genome versus it’s sequencing depth.

**Figure S2:**
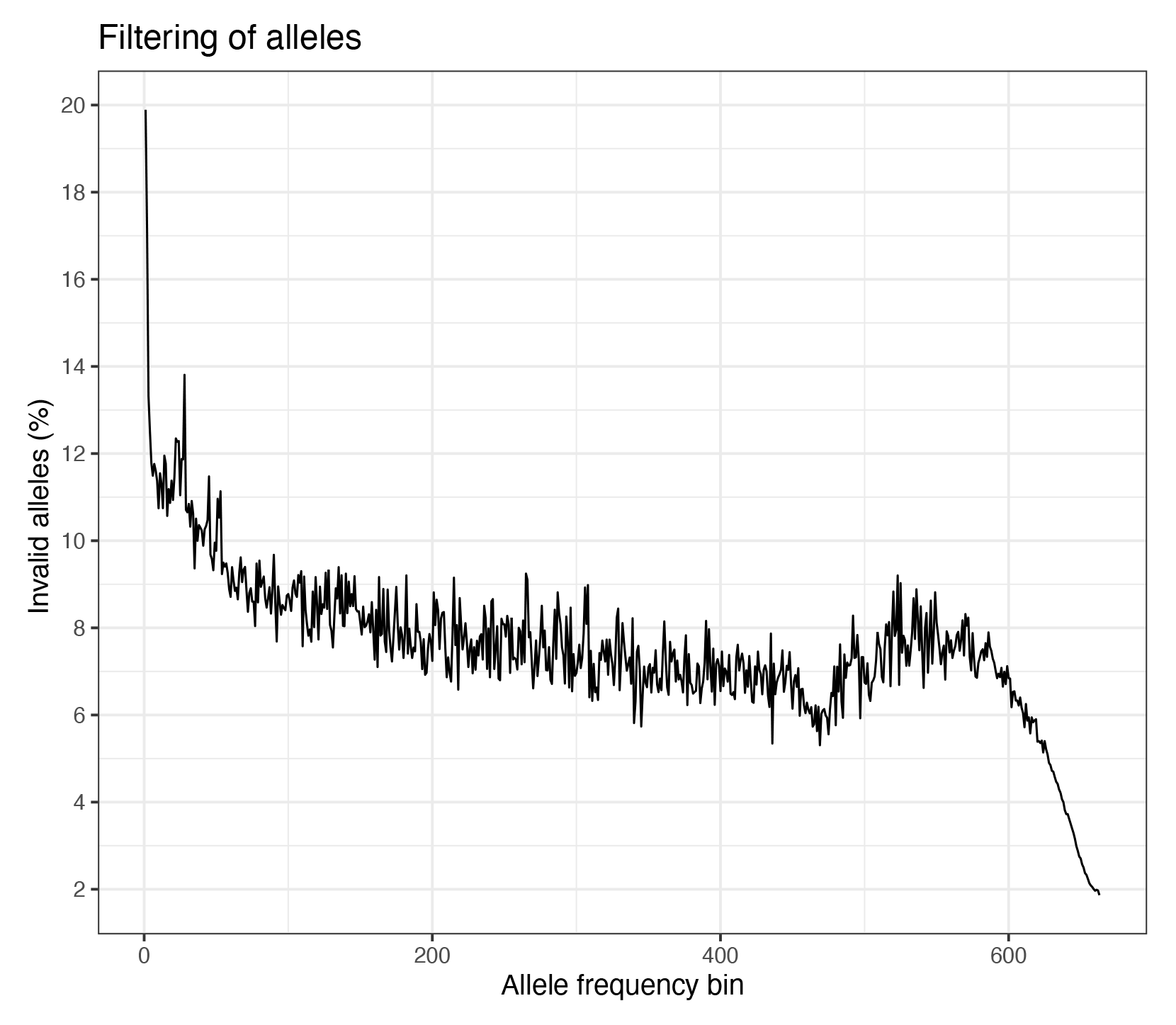
The proportion of alleles that failed validation and were discarded, stratified by allele frequency.

**Figure S3:**
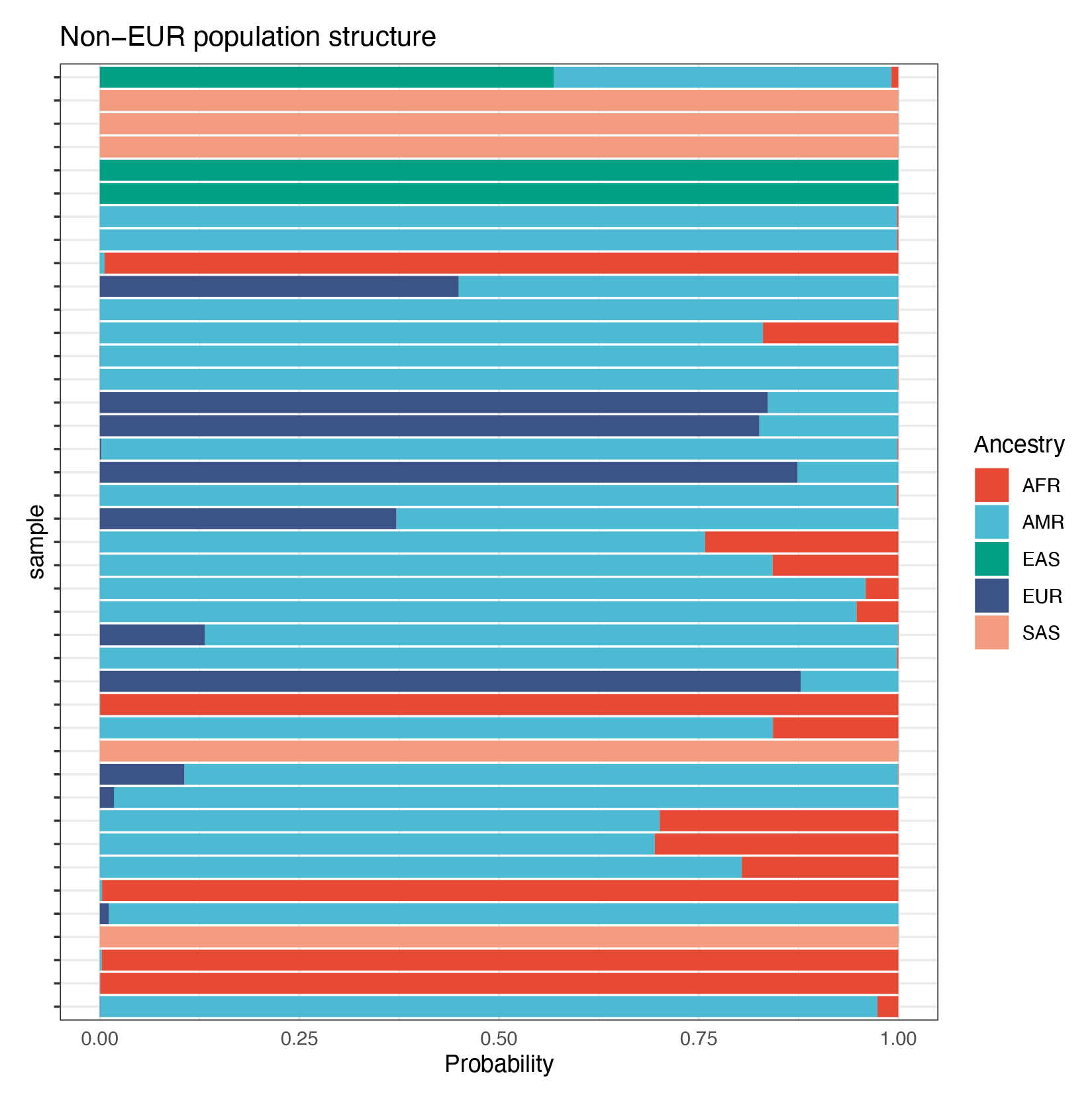
Genome ancestry mapping results with somalier based on on SNVs, for genomes with more than 10% EUR contribution.

**Figure S4:**
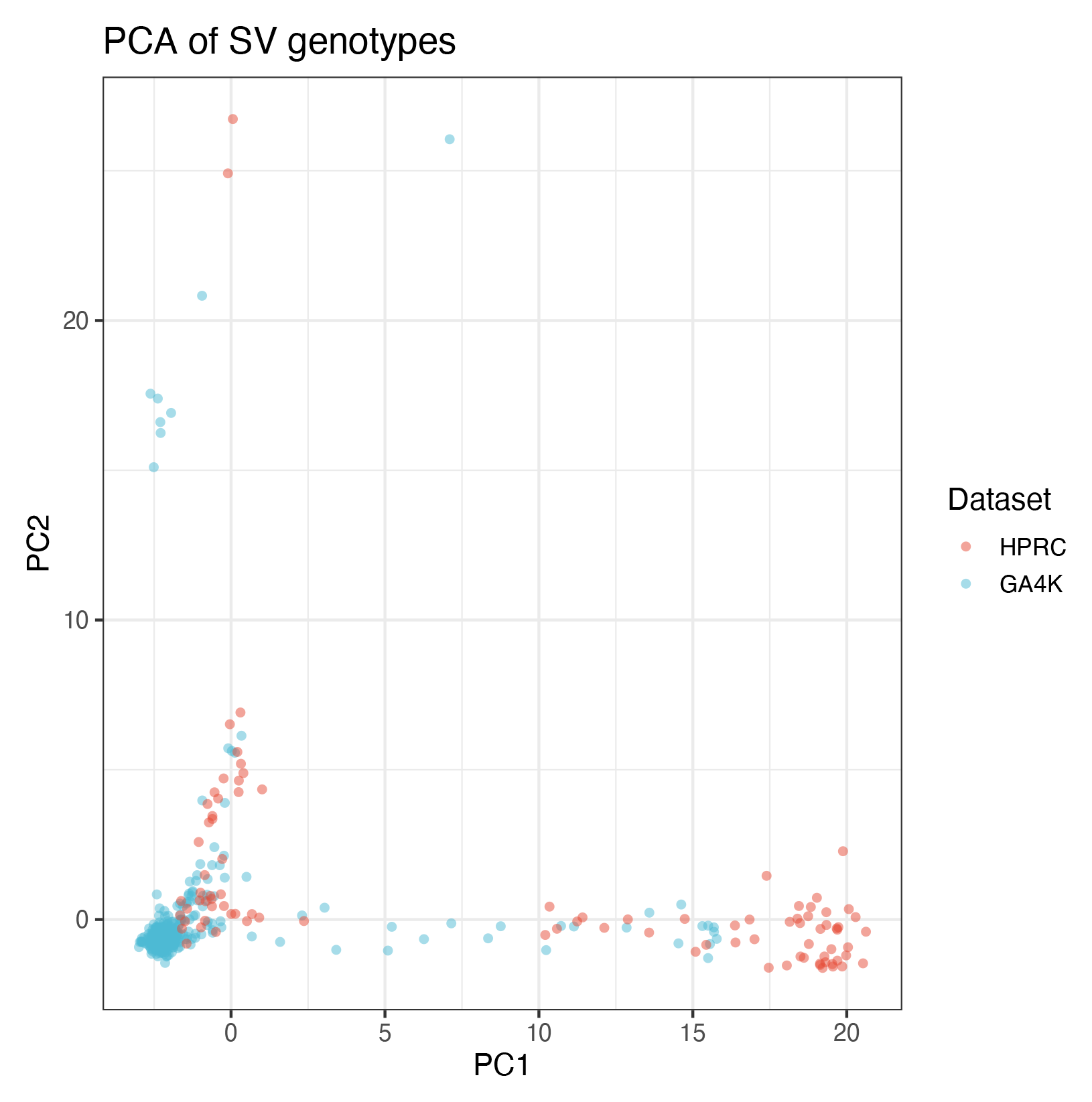
Population structure obtained using a PCA projection of the full set of GA4K and HPRC SVs that were called with minigraph.

**Figure S5:**
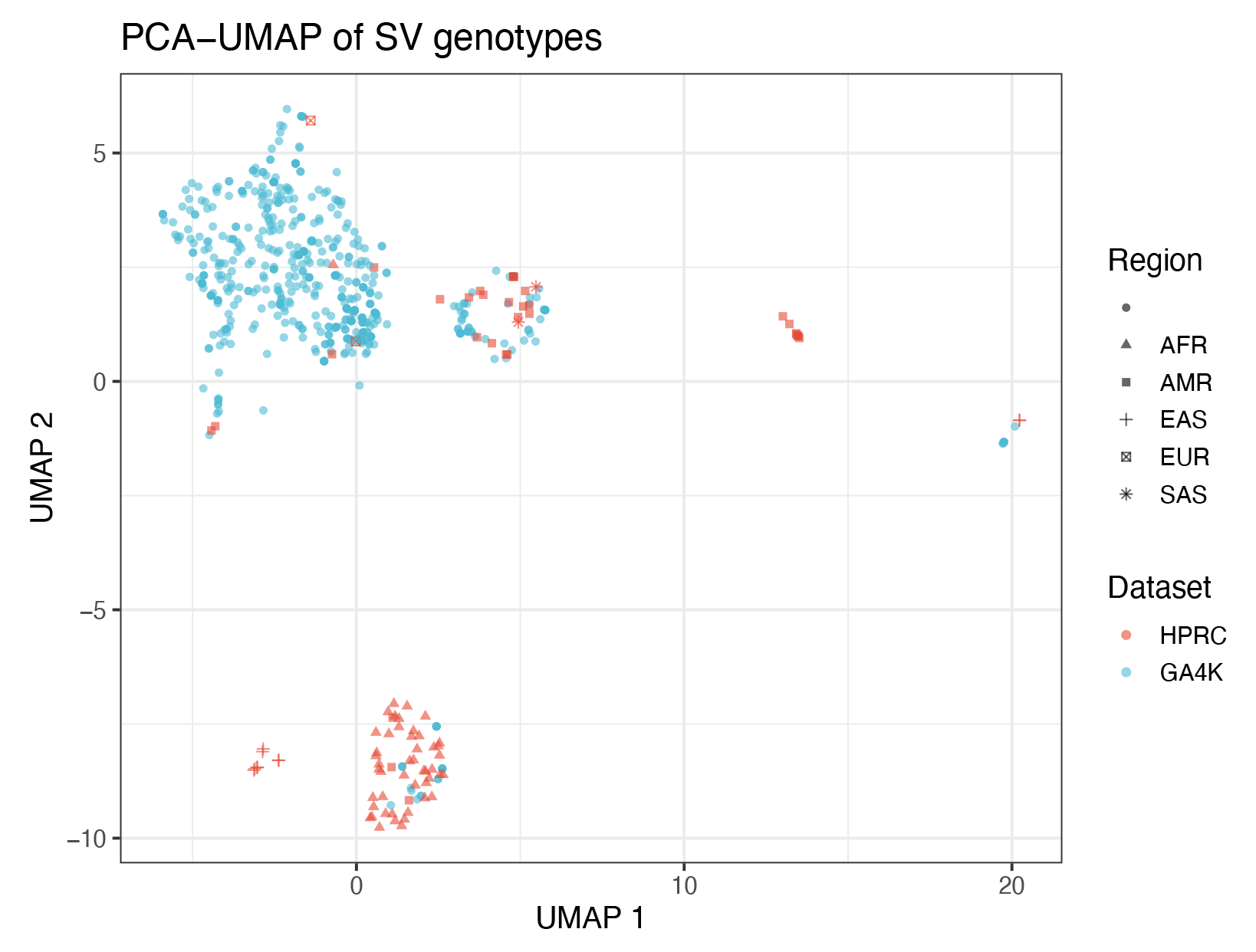
Population structure obtained using a PCA-UMAP projection that was trained on a subset of GA4K and HPRC SVs that were called with minigraph.

**Figure S6:**
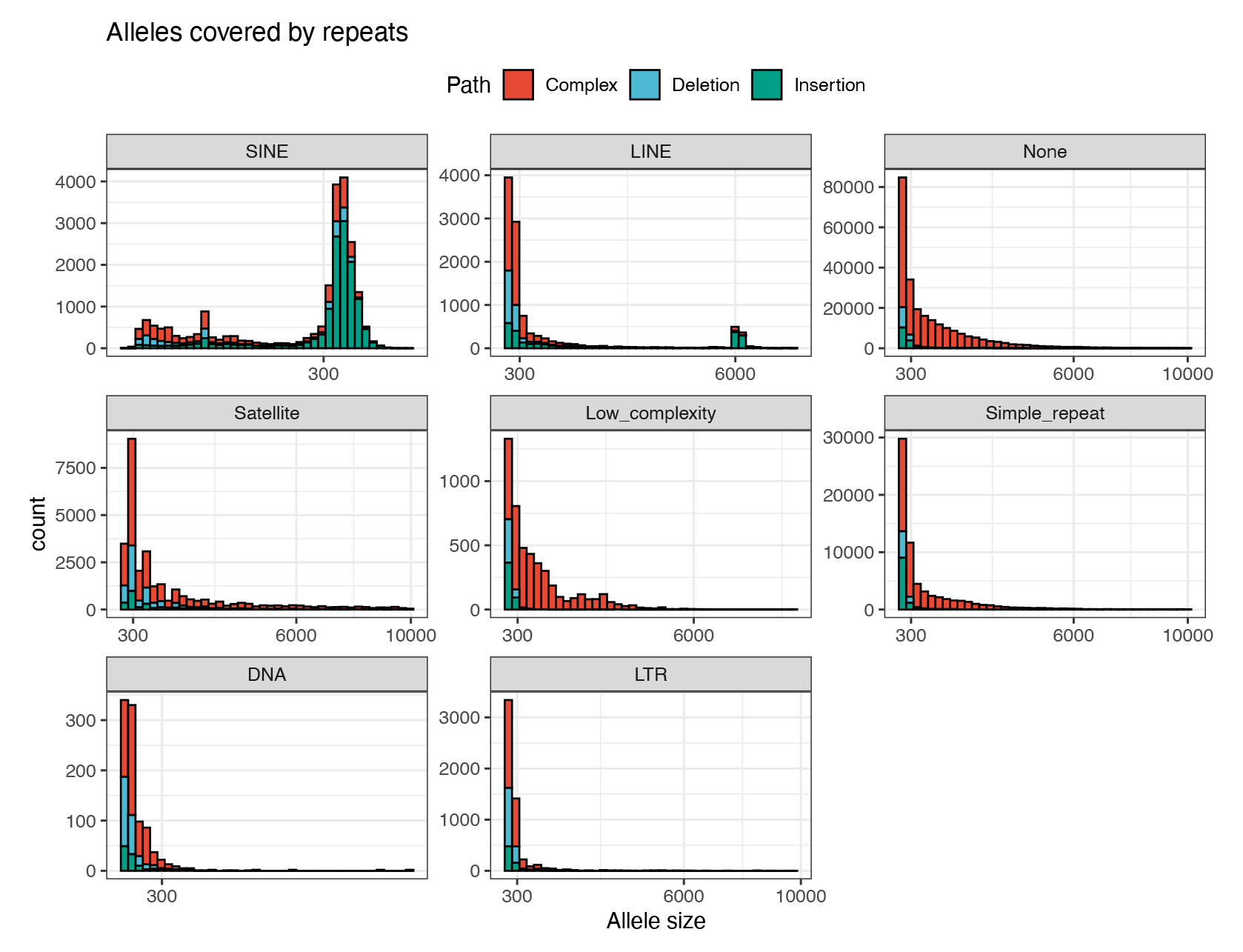
Repeat spectrum in allele sequences in the pangenome, masked with RepeatMasker and stratified by allele type. Only alleles that are 80% covered by the annotation over their span are shown.

**Figure S7:**
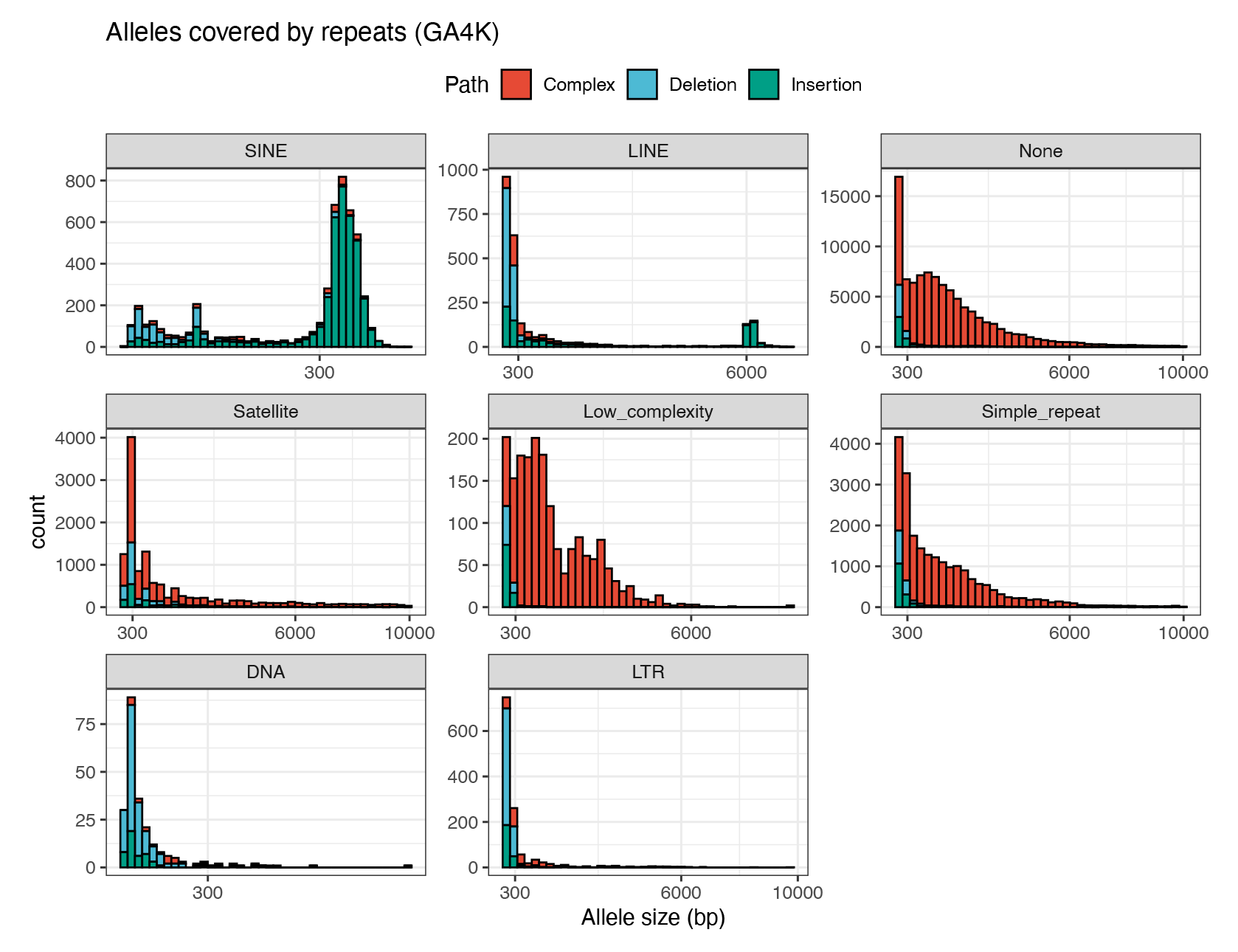
Repeat spectrum in allele sequences that are unique to GA4K, masked with RepeatMasker and stratified by allele type. Only alleles that are 80% covered by the annotation over their span are shown.

**Figure S8:**
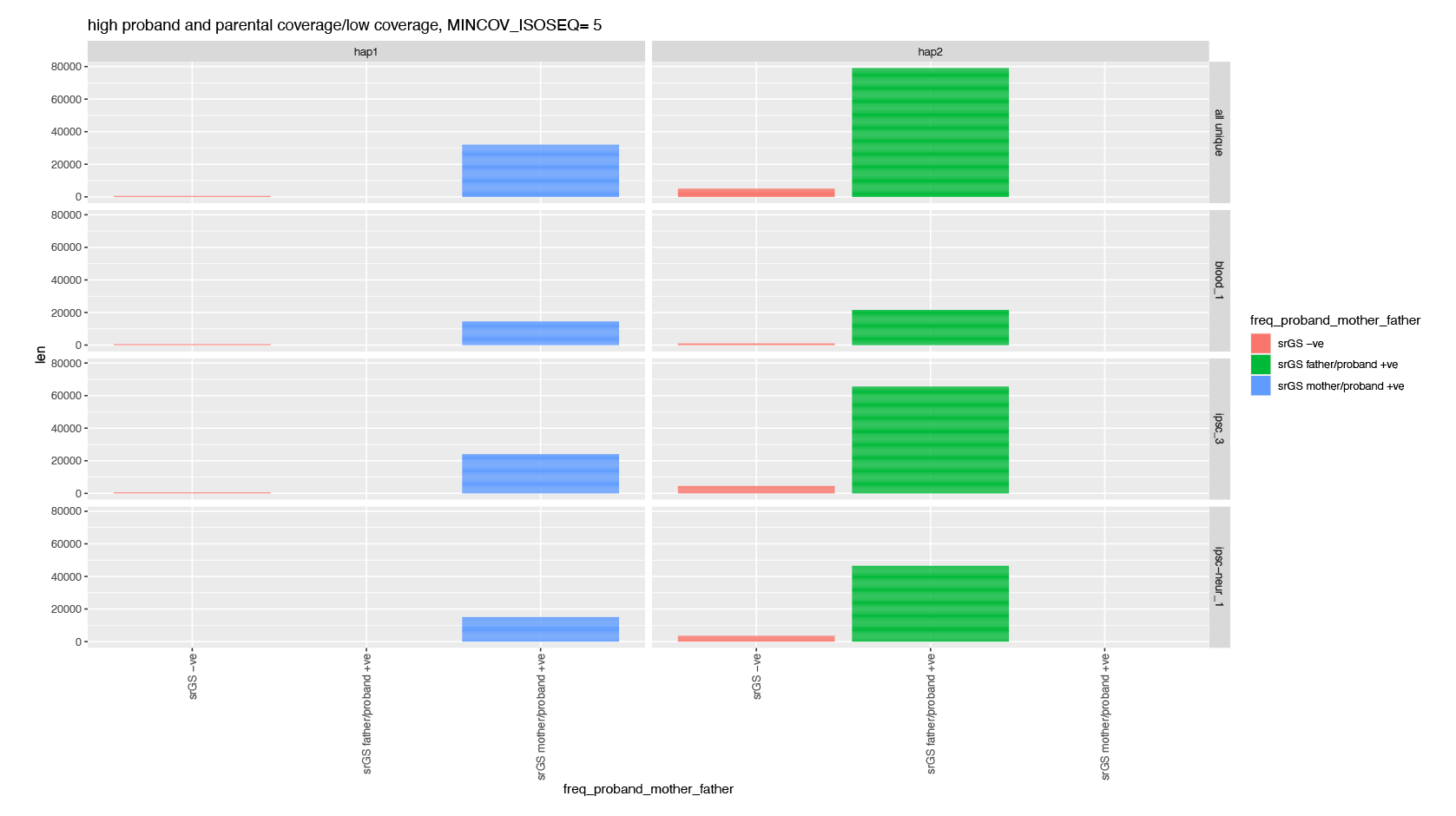
Span of sequence that is covered by short-reads and Iso-Seq reads in contigs that are in a proband’s assembly but that are not anchored in the genome graph.

**Figure S9:**
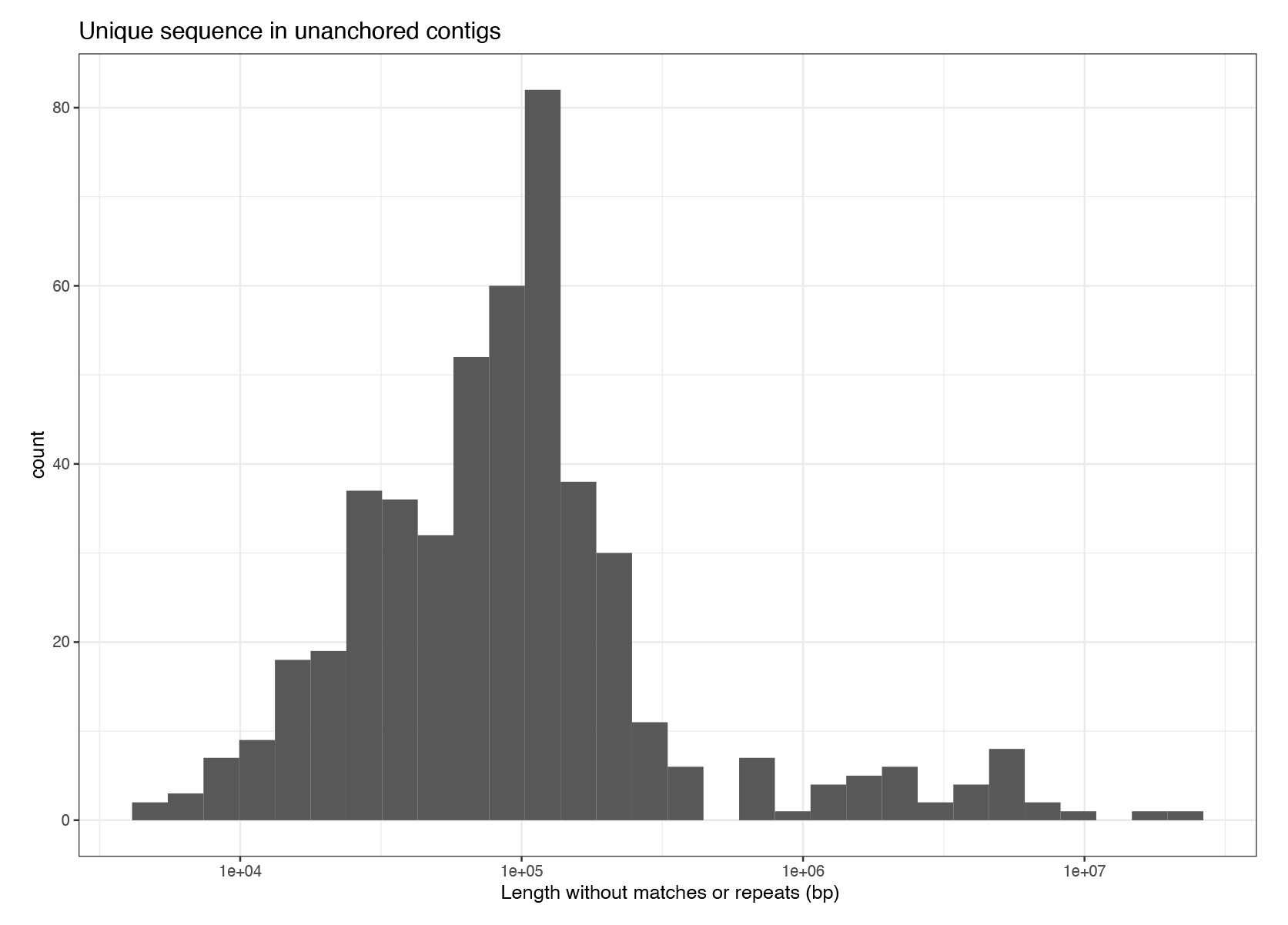
Amount of sequence in unanchored contigs in each genome that does not map to chm13v2.0 and is not covered by repeats.

**Figure S10:**
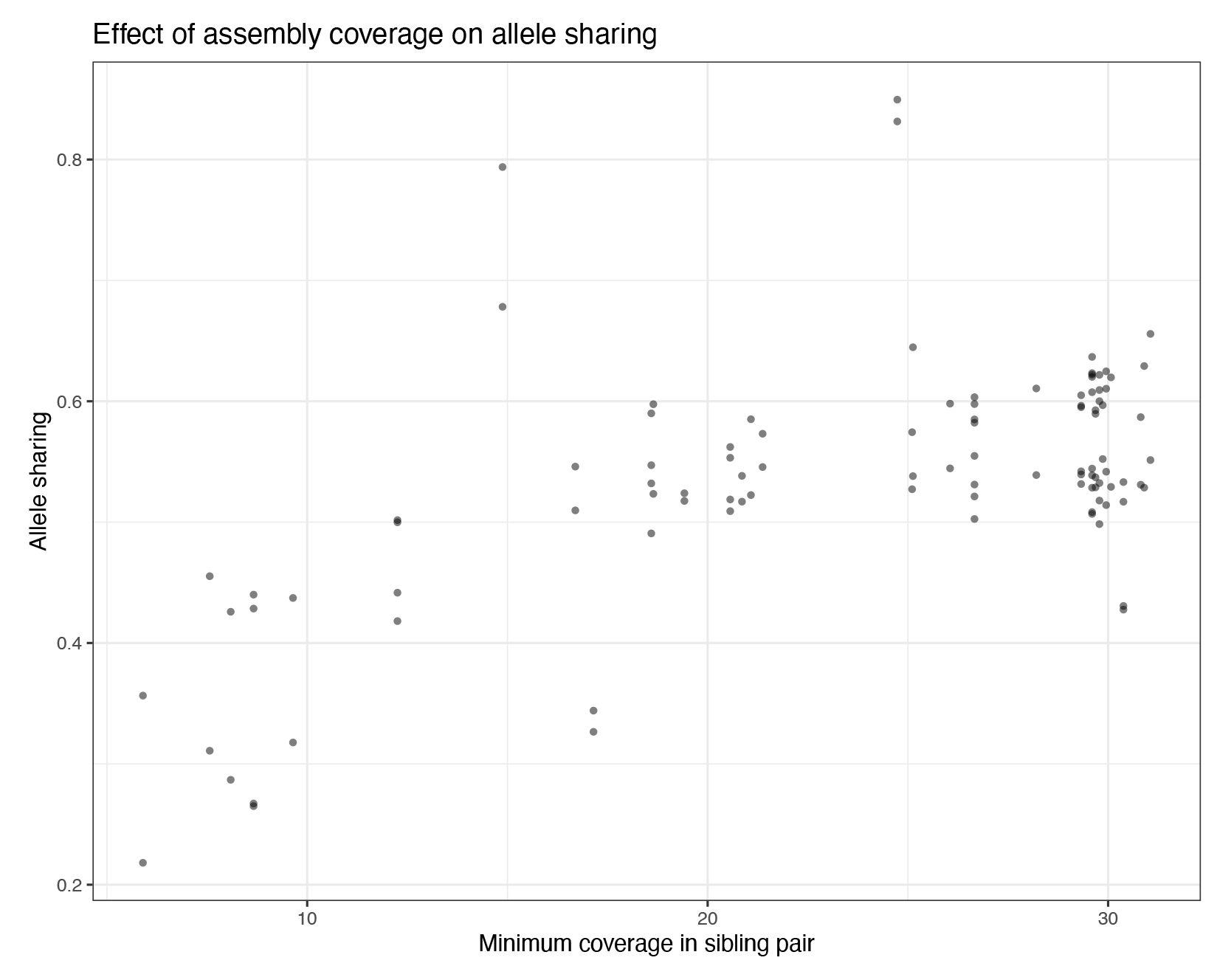
The proportion of allele sharing between pairs of siblings versus the lowest sequencing depth of the genome pair.

**Figure S11:**
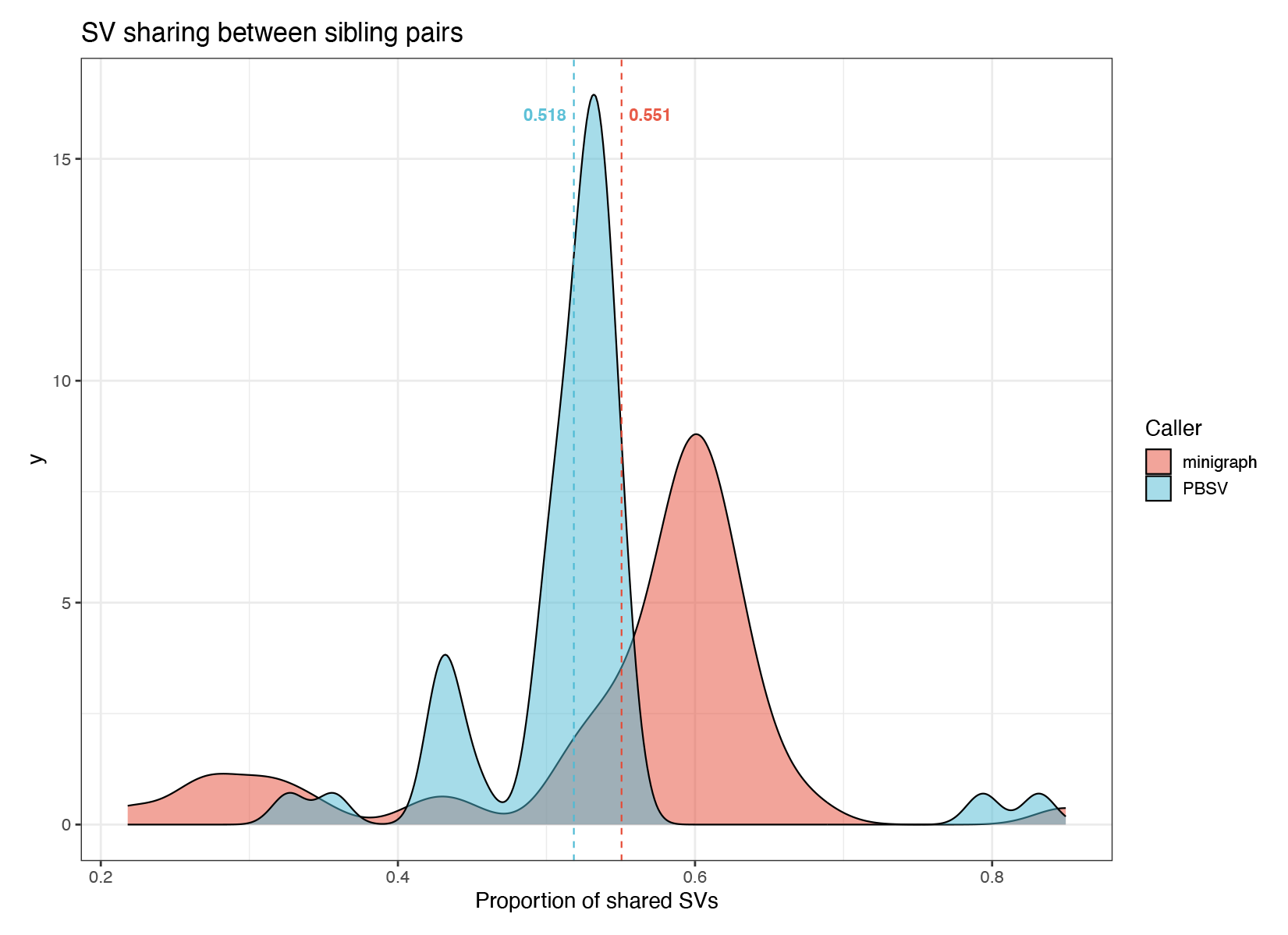
C) Allele sharing between siblings in GA4K (twins excluded and low coverage pairs included), calculated with PBSV versus minigraph.

**Figure S12:**
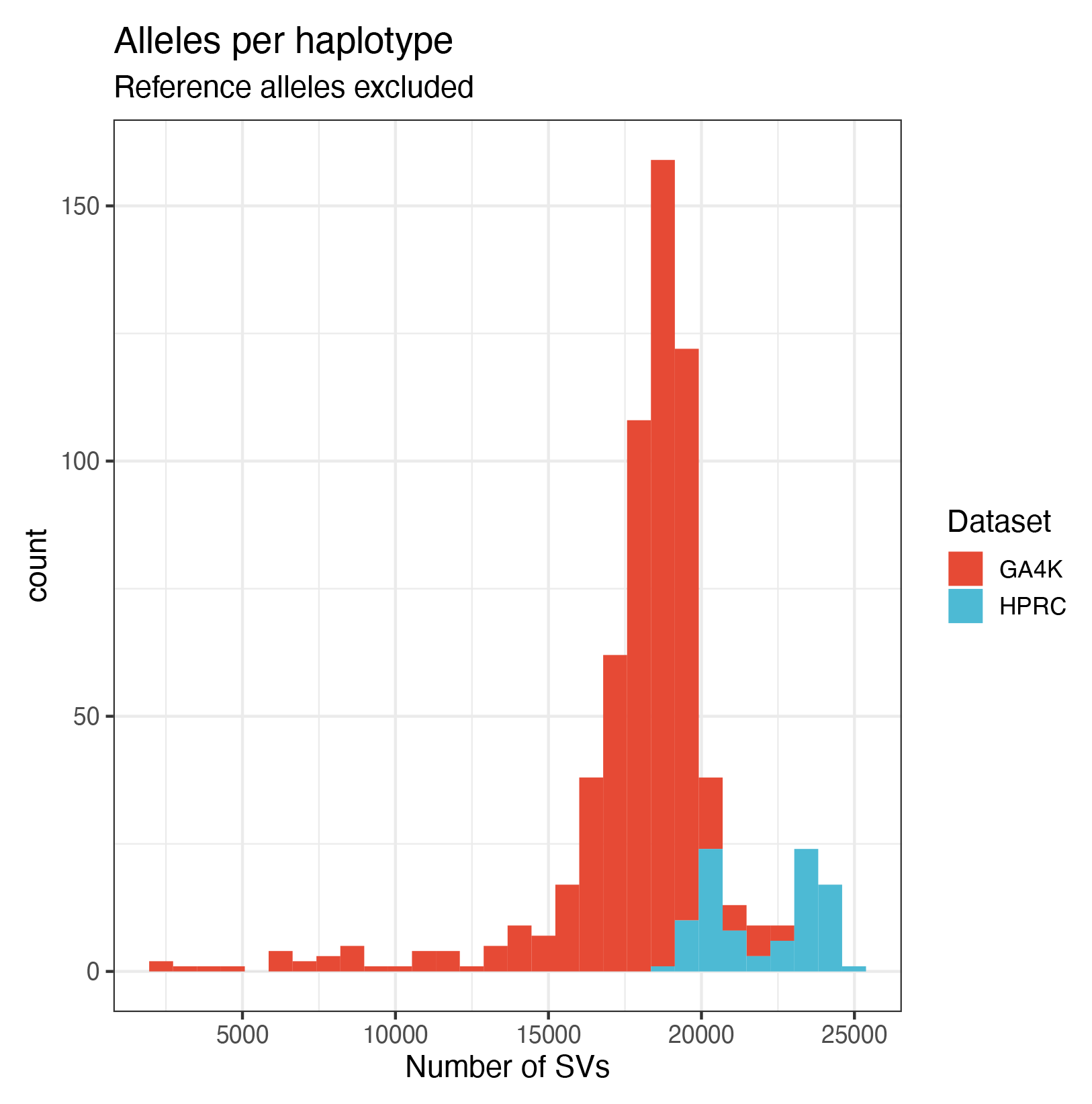
Distribution of non-reference alleles per genome, stratified by HPRC and GA4K cohorts.

**Figure S13:**
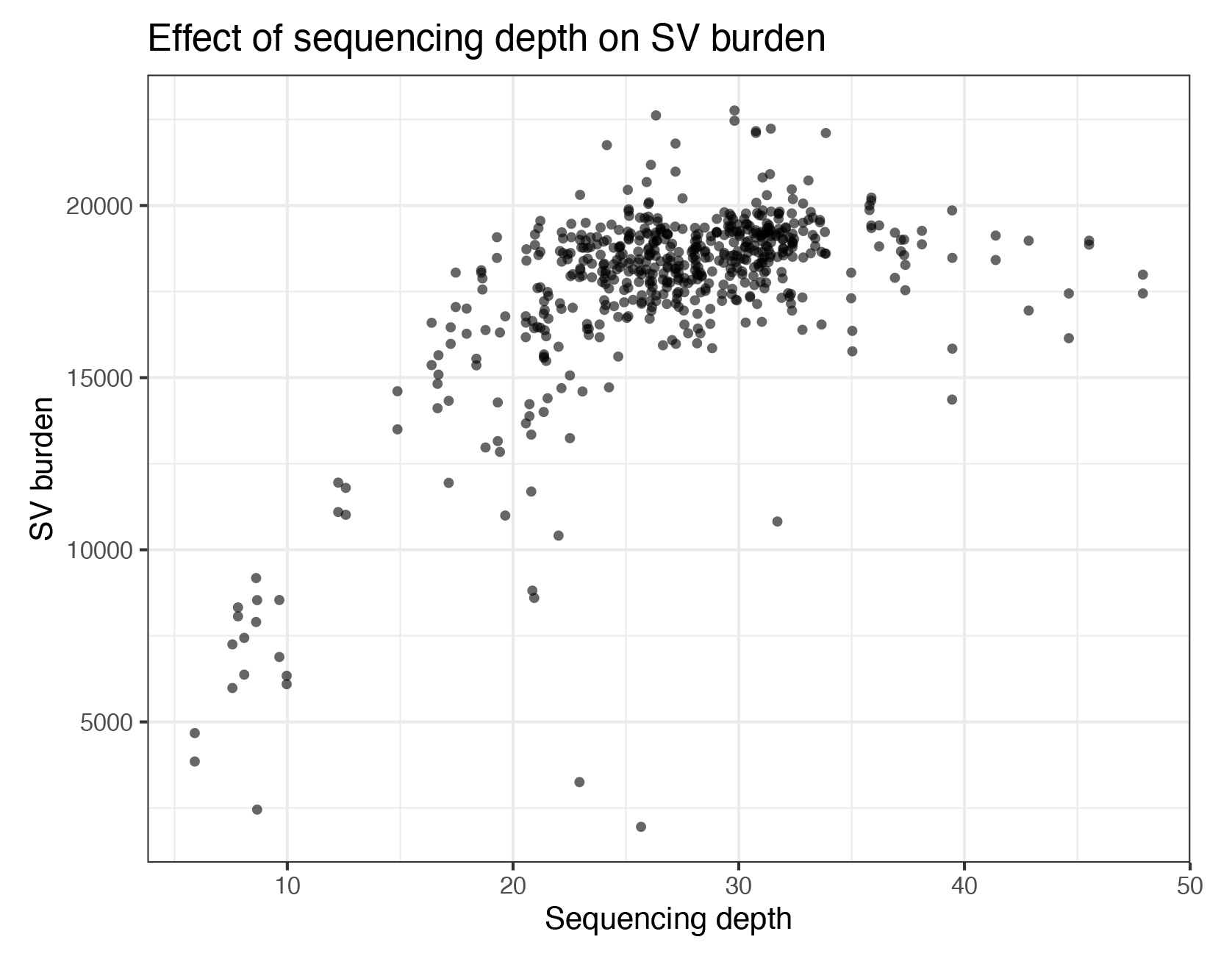
SV burden of each genome in GA4K vs the sequencing depth of the genome.

**Figure S14:**
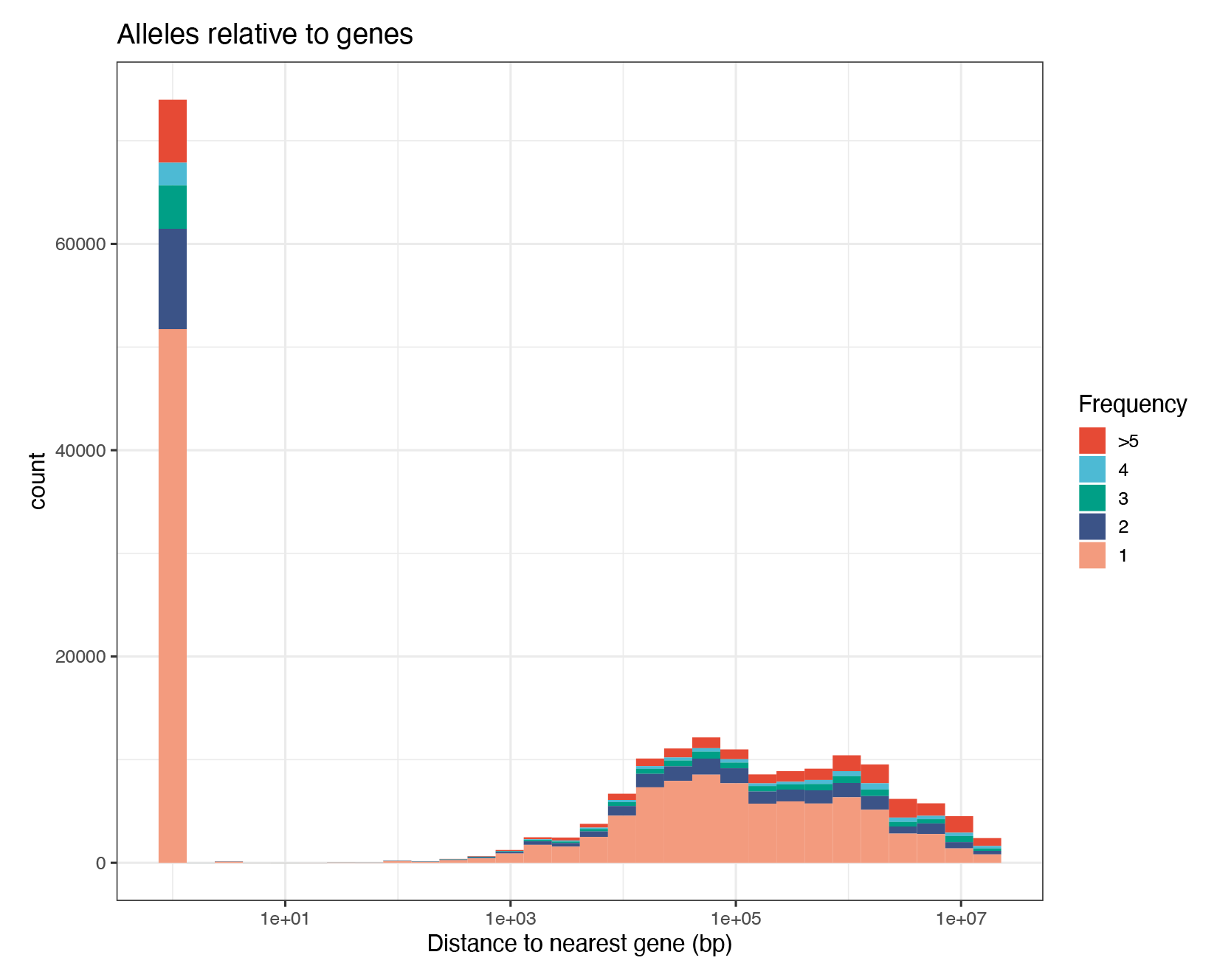
Distance distribution of SV alleles that are unique to GA4K from the nearest gene, stratified by allele frequency.

**Figure S15:**
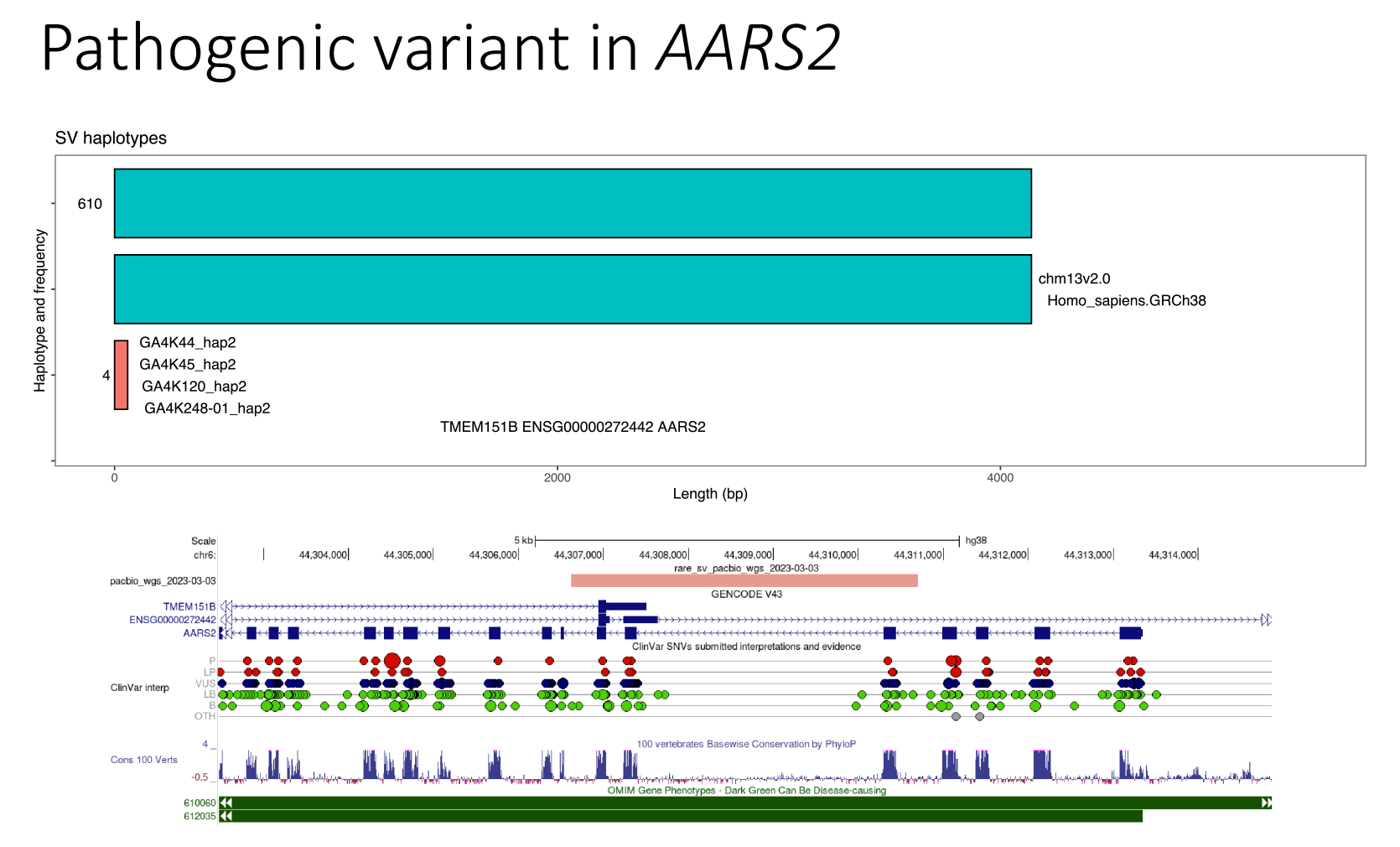
Inversion in the ACOX1 gene. Showing minigraph haplotype structures with frequencies. Each haplotype is an ordering of nodes. The minigraph bubble representing the inversion is also shown.

**Figure S16:**
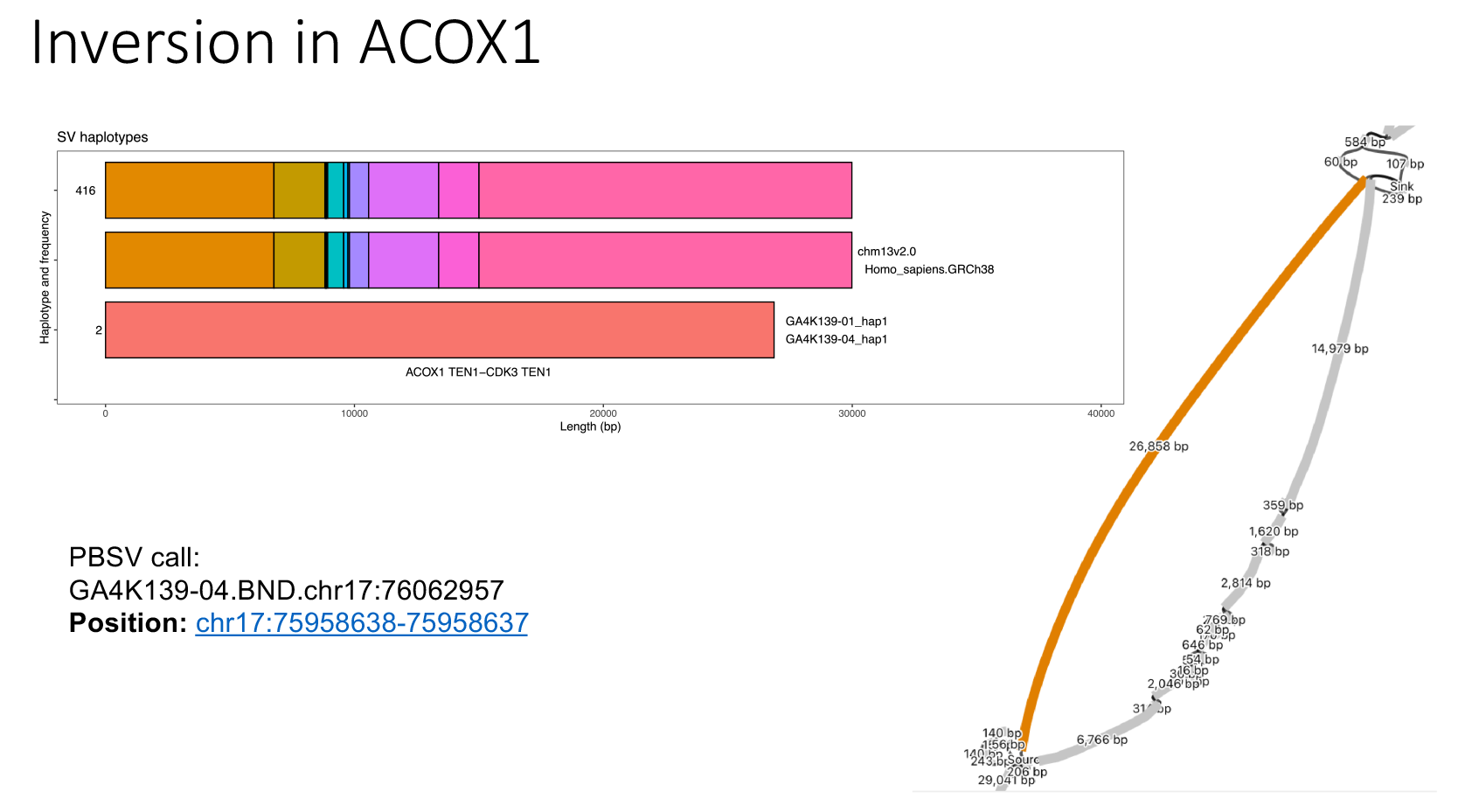
Pathogenic variant in AARS2. Showing minigraph haplotype structures with frequencies and the UCSC genome browser view of the over-lapping PBSV calls.

**Table S1:** Number of contigs in GA4K86-01 diploid assembly that are not anchored in the genome graph but that are covered by short reads originating from the trio.

| Mean bp | Maternal Assemblies | Paternal Assemblies |
| --- | --- | --- |
| Total Assembly Size | 3025967107.0 | 2899273161.5 |
| Total Unanchored Size | 17425226.0 | 26779214.5 |
| Short-read covered both parents | 16130019.0 | 23518471.5 |
| Short-read maternal-only | 952199.5 | 14706.5 |
| Short-read paternal-only | 43020.0 | 2572237.5 |

**Table S2:** Composition of SV concordant and discordant sets and the number of SVs recovered by lllumina Manta SV calls in the concordant/discordant sets (Methods).

| Rare (5%) PacBio/Illumina SVs, rare (5%) 2-allele MG calls |  |  |  |  |  |  |
| --- | --- | --- | --- | --- | --- | --- |
| MG alleles | PB SVs | ILMN SVs | Concordant SVs | Discordant SVs | Concordant rep # | Discordant rep # |
| 679 | 3064 | 4357 | 185 | 2879 | 112 (61%) | 229 (8%) |

| All PacBio/Illumina SVs, 2-allele MG calls |  |  |  |  |  |  |
| --- | --- | --- | --- | --- | --- | --- |
| MG alleles | PB SVs | ILMN SVs | Concordant SVs | Discordant SVs | Concordant rep # | Discordant rep # |
| 95594 | 22205 | 11610 | 5448 | 16757 | 3060 (56%) | 5503 (33%) |

**Table S3:** Summary of leading rare GA4K alleles in phrank rankings and that map to OMlM genes, together with their pathogenic potential.

| Chromosome | Start | End | Gene | Genotype | Phrank score | Sample | Description |
| --- | --- | --- | --- | --- | --- | --- | --- |
| chr2 | 73448340 | 73448766 | *ALMS1* | 0/1 | 10.734 | GA4K6-01 | carrier |
| chr16 | 1464967 | 1471483 | *CLCN7* | 0/1 | 21.714 | GA4K33 | DEL of EXON, LOF linked only to AR disease, no 2nd coding mt |
| chr2 | 168954038 | 168961552 | *ABCB11* | 1/0 | 5.955 | GA4K34 | carrier |
| chr12 | 68691995 | 68691995 | *NUP107* | 1/0 | 23.635 | GA4K42 | exon adjacent INS, AR disease, no 2nd mutation |
| chr14 | 92014358 | 92016430 | *TRIP11* | 0/1 | 7.491 | GA4K43 | carrier; (multi kb DEL with exons, inherited), no 2nd variant |
| chr6 | 44306619 | 44310759 | *AARS2* | 0/1 | 6.262 | GA4K45 | **diagnostic (2nd allele SNV)** |
| chr16 | 89747687 | 89797146 | *FANCA* | 1/1 | 11.644 | GA4K50 | LARGE 50kb GENE DEL inherited from 1 parent, no 2nd variant |
| chr16 | 46692790 | 46701252 | *ORC6* | 1/0 | 19.056 | GA4K87-01 | carrier; (multi kb DEL with exons, maternal), no 2nd variant |
| chr9 | 133062454 | 133083012 | *CEL* | 1/0 | 6.464 | GA4K87-01 | frameshift VNTR (200bp) in terminal exon - reported with uncertain significance in disease - no reported DM in family |
| chr1 | 202332934 | 202341602 | *UBE2T* | 0/1 | 36.288 | GA4K88-01 | carrier; (multi kb DEL with exons, paternally inherited), no 2nd variant |
| chr16 | 50722536 | 50723555 | *NOD2* | 1/0 | 11.684 | GA4K90-01 | DEL of NOD2 where missense variation linked to BLAU SDR - low phenotypic fit |
| chr22 | 17183464 | 17189024 | *ADA2* | 1/0 | 10.841 | GA4K93-01 | carrier |
| chr14 | 87925160 | 87956830 | *GALC* | 0/1 | 8.468 | GA4K105-01 | carrier; DEL of GALC reported Likely Pathogenic in ClinVar, AR disease, no 2nd coding mt |
| chr7 | 138095272 | 138099856 | *AKR1D1* | 0/1 | 7.247 | GA4K116-01 | carrier |
| chr17 | 18293722 | 18356461 | *TOP3A* | 0/1 | 5.407 | GA4K124-01 | carrier; (60kb DEL with exons + other genes, paternal), AR disease - no 2nd variant |
| chr2 | 162280989 | 162282198 | *IFIH1* | 0/1 | 6.171 | GA4K137-01 | DEL of IFIH1 exon, immunodeficiency linked to AR LOF, not fitting pt phenotype |
| chr19 | 53805788 | 53808020 | *NLRP12* | 0/1 | 19.475 | GA4K153-01 | DEL of exon where LOF SNV is ClinVar LP variant in *NLRP12* causing AD cold autoinflammatory sdr; phenotypic FIT? DIAGNOSTIC? |
| chr2 | 231272533 | 231275041 | *ARMC9* | 0/1 | 7.184 | GA4K157-01 | carrier |
| chr8 | 142875528 | 142913101 | *CYP11B1* | 1/0 | 13.311 | GA4K158-01 | carrier; DEL of CYP11B1 repoted Pathogenic in ClinVar, LOF linked only to AR disease, no 2nd coding mt |
| chr7 | 107313633 | 107328880 | *COG5* | 1/0 | 16.803 | GA4K166-01 | carrier; (15kb DEL with exon, maternal), no 2nd variant |
| chr17 | 11784064 | 11784935 | *DNAH9* | 0/1 | 20.352 | GA4K171-01 | carrier; DEL of EXON, LOF linked to AR disease, no 2nd coding mt |
| chr9 | 36247443 | 36254553 | *GNE* | 1/0 | 43.179 | GA4K171-01 | carrier; DEL of GNE exon, maternally inherited, no 2nd coding mt |
| chr7 | 105074927 | 105089397 | *KMT2E* | 1/0 | 8.099 | GA4K178-01 | Exonic DEL in gene where LOF causes AD disease **inherited from partially affected mom -**<br>**DIAGNOSTIC** |
| chr12 | 56044149 | 56044149 | *RPS26* | 1/0 | 9.296 | GA4K180-01 | 3'UTR DUP (inherited from unaffected mother) |
| chr17 | 75958823 | 75988811 | *ACOX1* | 0/1 | 9.3 | GA4K196-01 | for follow-up; large INV variant involving 5' exons: gene with gain of function mutations, requires RNA studies |
| chr16 | 78150417 | 78229469 | *WWOX* | 1/0 | 10.155 | GA4K223-01 | carrier; (75kb DEL with exon, maternal), AR disease - no 2nd variant |
| chr9 | 34645665 | 34651238 | *GALT* | 1/0 | 12.818 | GA4K231-01 | carrier; DEL of GALT, AR disease, no 2nd coding mt |
| chr12 | 76334357 | 76393510 | *BBS10* | 1/0 | 6.119 | GA4K234-01 | carrier |
| chr7 | 152659511 | 152660903 | *XRCC2* | 0/1 | 6.672 | GA4K237-04 | carrier; (1.3 kb DEL with exon, maternally inherited), no 2nd variant |
| chr6 | 44306619 | 44310759 | *AARS2* | 0/1 | 6.738 | GA4K243-01 | carrier |

## Notes

### Competing Interest Statement

Juniper Lake is a current or past employee of Pacific Biosciences.

### Funding Statement

This work was made possible by the generous gifts to Children's Mercy Research Institute and Genomic Answers for Kids program at Children's Mercy Kansas City.

### Author Declarations

The Institutional Review Board (IRB) of Children's Mercy Kansas City gave ethical approval for this work (Study#11120514).

